# SARS-CoV-2 Spread Under the Controlled-Distancing Model of Rio Grande do Sul, Brazil

**DOI:** 10.1101/2023.06.09.23291044

**Authors:** Ricardo Rohweder, Lavínia Schüler-Faccini, Gonçalo Ferraz

## Abstract

In early 2020, the government of Rio Grande do Sul established a public-health assessment-response framework to halt the spread of SARS-CoV-2, called ‘controlled-distancing model’ (CDM). This framework subdivided the state in 21 regions where it evaluated a composite index of disease transmission and health-service capacity. Updated on a weekly basis, the index placed regions on a color-coded scale of flags, which guided adoption of non-pharmaceutical interventions. We aim to evaluate the extent to which the CDM accurately assessed transmission and the effectiveness of its responses throughout 2020. We estimated the weekly effective reproduction number (*R*_*t*_) of SARS-CoV-2, for each region, using a renewal-equation-based statistical model of notified COVID-19 deaths. Using *R*_*t*_ estimates, we explored whether flag colors assigned by the CDM either reflected or affected SARS-CoV-2 dissemination. Flag assignments did reflect variations in *R*_*t*_, to a limited extent, but we found no evidence that they affected R_t_ in the short term. Medium-term effects were apparent in only four regions after eight or more weeks of red-flag assignment. Analysis of Google movement metrics showed no evidence that people moved differently under different flags. The dissociation between flag colors and the propagation of SARS-CoV-2 does not support the claim that non-pharmaceutical interventions are ineffective. Our results show, however, that decisions made under the CDM framework were ineffective both for influencing the movement of people and for halting the spread of the virus.

## Introduction

Brazil is home to less than 3% of the world population but had more than 10% of the global number of COVID-19 deaths.^1^ For the first weeks of the pandemic, the average number of secondary infections originating from one infected host—or *R*_*t*_, the effective reproduction number—was higher than 3 in São Paulo and Rio de Janeiro.^2^ By late March, estimated *R*_*t*_ decreased sharply,^2^ following implementation of the first non-pharmaceutical interventions (NPIs), but the weekly nationwide number of cases and deaths continued to increase until early August, 2020.^1^ In the absence of a vaccine or effective treatment, the only way of keeping *R*_*t*_ consistently below 1 was to improve NPI implementation with good decisions about when and how to restrict travel, schools, commerce and other forms of activity. Rio Grande do Sul (RS), which declared a state of calamity on March 19, 2020, was the first Brazilian state to attempt a public-health assessment-response framework for the implementation of NPIs. This framework, established May 10, and called *Modelo de Distanciamento Controlado* (in English, Controlled Distancing Model, henceforth CDM),^3^ was in force throughout the remainder of 2020 and is the focus of our study.

The COVID-19 pandemic posed a decision-making challenge to public health authorities around the world.^4^ NPI implementation embodies the three core elements of any structured decision: what one wants, what one knows, and what one can do. ^5^ The range of possible interventions captures what one can do; the rate of pathogen transmission is what ought to be known; and stopping dissemination of a disease is, hopefully, what one wants. From its inception, however, the CDM reflected the false dichotomy between protecting lives and protecting livelihoods,^6^ which influenced municipal, state, and federal policies in Brazil throughout the pandemic. Thus, the RS state government (henceforth, the government) presented the CDM as a ‘mixed, modular, mutually-agreed strategy for balancing the priority of life with economic recovery’ (our translation).^7^ The framework was rooted on a color-coded scale of flags (yellow, orange, red, black) assigned on a weekly basis to each of 21 COVID regions. A region’s flag color was given by an *ad hoc* combination of eleven metrics grouped in two sets (Table S1), which carried equal weight on a composite index.^3^ One set assessed the pace of disease transmission; another focused on healthcare service capacity to assist COVID patients. Each color specified a range of NPIs applicable to different segments of human activity with varying levels of restrictiveness, depending on the segment’s putative contribution for spreading the virus and relevance to the state’s economy. The correspondence between flag colors and NPIs was frequently revised throughout the CDM implementation, with municipalities appealing flag assignments that they deemed too harsh and occasionally obtaining less restrictive colors. The CDM was in place until May 15, 2021, when the government replaced it by an even more flexible assessment-response framework (Table S2).^8^

The COVID-19 pandemic strengthened a rich literature about the effectiveness of NPIs. Early on, Dehning et al.^9^ showed that a contact ban and closure of non-essential commerce helped Germany control the spread of the virus. Flaxman et al.,^10^ using data from eleven European countries, demonstrated the effectiveness of lockdowns for reducing *R*_*t*_. Li et al.,^11^ with a sample of 131 countries, found that *R*_*t*_ decreased after school closures, workplace closures, and public event bans. These analyses help decision makers chose among possible interventions. Frameworks like the CDM, however, require not only knowledge about what interventions work best, but also good decisions about timing and extent of intervention. CDM flag assignments convey timing, while the measures associated with each flag convey extent decisions, embodied by NPI choice. The combined result of timing and NPI choice determines the effectiveness of an assessment-response framework.

We assess the effectiveness of the RS CDM throughout 2020 focusing on two central questions about flag assignment and its relationship with *R*_*t*_. First, did *R*_*t*_ respond to flag assignments, as it should when interventions produce the desired result? And second, did flag assignment reflect previous changes in *R*_*t*_, as expected from an adequate assessment of disease transmission? To pursue these questions, we obtained weekly estimates of *R*_*t*_ for each RS COVID region and contrasted estimates with corresponding flag colors. Because subnotification of disease cases was unavoidable, we employed a statistical model of *R*_*t*_ informed by the number of notified COVID-19 deaths, an imperfect but more reliable data source than the number of notified cases.^10,12,13^ Our longitudinal assessment of the effectiveness of the CDM stops at the end of 2020 because the beginning of 2021 saw the arrival of a new, more transmissible SARS-CoV-2 lineage, and shortly afterward, the termination of the CDM.

## Methods

### Data

Our study combines epidemiologic, demographic, mobility, and assessment-response data collected between 15 March and 21 December, 2020, in the 21 RS COVID regions. Daily reported infections and deaths were obtained from the *Painel Coronavírus do Ministério da Saúde*.^14^ Projected human population age structure per municipality, came from the *Fundação de Economia e Estatística do Rio Grande do Sul*^15^ and municipality-level mobility metrics from Google’s COVID-19 Community Mobility Reports.^16^ Google provides metrics of change in daily mobility with reference to January 2020 for six place categories: Residential, Transit stations, Parks, Grocery & pharmacy, Retail & recreation, and Workplaces. We computed region-level mobility metrics as arithmetic means of the respective municipality values weighted by each municipality’s population size. Weekly, region-specific assessment-response data—the CDM flag colors—were obtained from the government website.^17^ Codes and processed data are available as supplementary material to this paper and in an online repository.^18^

### Epidemiological Model

The core of our analytical approach is an adaptation of Flaxman et al.’s^10^ model, which uses a renewal equation to infer the latent daily number of SARS-CoV-2 infections that result in a reported number of COVID-19 deaths. Such inference entails the estimation of the effective reproduction number (*R*_*it*_, or *R*_*t*_, for short) and attack rate (*p*_*it*_; proportion of the population infected) for every RS COVID-19 region *i* and day *t*. The estimation of the true, partially observed, number of cases from the observed number of deaths draws on a vector of infection fatality ratios and three distributions obtained from the literature.

Infection fatality ratios *f*_*i*_ were adjusted to the population age-structure of each region while considering the frequency of contacts among age classes summarized by a contact matrix estimated by Grijalva et al.^19^ (Supplementary Material Methods and Code 1). The three distributions, which quantify the temporal connection between deaths and cases were: 1) a distribution of time from the onset of symptoms to death obtained by Ranzani et al.^20^ from Brazilian data, with mean 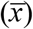 of 15.4 and standard deviation (*s*) of 6.9 days; 2) a distribution of times from infection to onset of symptoms (or incubation time) estimated by Linton et al.^21^ and Li et al.^22^ at the beginning of the pandemic, with 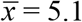 and *s* = 4.4 days; and 3) a distribution for the serial interval, the average time between a host’s infection and its transmission of the pathogen to another host, proposed by Flaxman et al.^10^ for SARS-CoV-2 spread in Europe, with 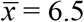 and *s* = 4.0 days. All three were modeled as gamma distributions with shape parameter 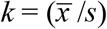 and scale parameter 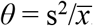. Based on these distributions and *f*_*i*_ values, we also model the true number of deaths. Our *R*_*t*_ estimates, furthermore, are related to the basic reproduction rate (*R*_0_) and the Google Mobility metrics as:

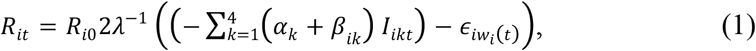

where *R*_*i*0_ is a basic reproduction number estimated for each COVID region *i* based on the data and a normal prior with mean 3.28^23^ and standard deviation 0.5. The notation *λ*^−1^ denotes the inverse logistic function, with coefficients *α*_*k*_ and *β*_*ik*_ representing, respectively, the effect of mobility metric *k* shared by all COVID regions, and the effect of mobility metric *k* specific to each COVID region. *I*_*ikt*_ is the value of the *k*’th mobility metric for region *i* on day *t*. Parameter 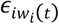 represents weekly autoregressive variation in region *i* that is not explained by variation in mobility. The mobility index *k* varies from 1 to 4 because we average the last three, highly correlated, metrics into one. The combined metrics refer to mobility in Grocery & pharmacy, Retail & recreation, and Workplaces. We processed data in the R^24^ framework and fit the model to data using an MCMC algorithm implemented in Stan^25^.

### CDM versus R_t_

To evaluate the relationship between *R*_*t*_ estimates and CDM flags we derived quantities of interest from the MCMC samples of the posterior distributions of epidemiological model parameters. Since flags were updated weekly and our model estimated daily *R*_*t*_’s, we first derived weekly *R*_*t*_ estimates as geometric means of daily *R*_*t*_’s, for every region. To examine whether flag changes associated with simultaneous or subsequent changes in *R*_*t*_, we compared distributions of *R*_*t*_ ratio (*R*_*t*+1_/*R*_*t*_) when flag restrictiveness had been downgraded, unchanged, or upgraded, respectively, on week *t* and *t*+1. To examine whether increases in *R*_*t*_ associated with subsequent upgrades in flag restrictiveness, we fit logistic models of flag upgrade probability as a function of *R*_*t*_ ratio, using lags of 1 to 7 weeks between ratio numerator and flag assignment. For each lag, we obtained a distribution of 4000 logistic slope parameters, each coming from one MCMC sample. We also compared the posterior distributions of *R*_*t*_ ratio among flag colors on week *t*, and plotted the relationship between duration of uninterrupted red-flag periods and the corresponding period’s *R*_*t*_ ratio (see Supplementary Material Methods and Code 2). To analyze the relationship between CDM flags and Google mobility metrics we built a mixed-effects model^26^ that accounts for region and temporal effects on mobility, described in Supplementary Material Methods and Code 3.

### Ethical Issues/Statement

This research was submitted to Plataforma Brasil, and approved by the Ethics Committee 5337 – Santa Casa da Misericórdia de Pelotas under the Certificate of Presentation of Ethical Appreciation (CAAE) 34335920.1.0000.5337 on October 9, 2020. This research was conducted without access to any individual information.

## Results

RS had 8,162 notified deaths due to COVID-19, and 409,794 notified cases of SARS-CoV-2 infection during our study period. Estimated attack rates (Table 1) ranged from 3.3% in Cachoeira do Sul to approximately 20% in Novo Hamburgo. Like Novo Hamburgo, the second, third, fourth and fifth COVID regions with the highest attack rates are in the Porto Alegre metropolitan area. More than half the regions had estimated attack rates above 10%.

**Table 1.** Total notified deaths and selected parameter estimates per COVID region, including estimated date of 10^th^ death, number of deaths, number of deaths per million inhabitants, number of infections, and attack rate between March 15 and December 21, 2020. Estimated deaths, infections and attack rates are show by their posterior distribution mean plus or minus one standard deviation (±σ). Regions marked with ‘*’ belong to the Porto Alegre metropolitan area.

| COVID region | Notified deaths | Date of 10<sup>th</sup> death | Deaths $\pm \sigma$ | Deaths per million | Infections (thousands $\pm \sigma$) | Attack rate % $\pm \sigma$ |
| --- | --- | --- | --- | --- | --- | --- |
| Porto Alegre* | 2 562 | April 10 | 2 590±121.0 | 1090 | 417.0±49.2 | 17.6±2.1 |
| Canoas* | 780 | May 28 | 789±45.0 | 1010 | 154.0±19.7 | 19.8±2.5 |
| Novo Hamburgo* | 775 | July 6 | 784±44.2 | 945 | 167.0±21.5 | 20.1±2.6 |
| Caxias do Sul | 715 | May 4 | 722±41.0 | 588 | 140.0±19.6 | 11.4±1.6 |
| Pelotas | 523 | June 24 | 537±35.1 | 611 | 92.8±14.1 | 10.6±1.6 |
| Passo Fundo | 486 | April 22 | 491±30.6 | 737 | 90.4±13.8 | 13.6±2.1 |
| Capão da Canoa* | 335 | June 19 | 339±25.0 | 854 | 63.2±10.2 | 15.9±2.6 |
| Guaíba* | 304 | June 19 | 310±22.4 | 750 | 55.0±7.7 | 13.3±1.9 |
| Santa Maria | 226 | June 8 | 228±18.7 | 407 | 36.4±5.9 | 6.5±1.1 |
| Uruguaiana | 191 | June 23 | 191±17.2 | 416 | 38.5±7.0 | 8.4±1.5 |
| Taquara* | 183 | July 12 | 187±17.3 | 797 | 36.9±5.6 | 15.7±2.4 |
| Lajeado | 180 | May 5 | 183±15.9 | 513 | 28.4±4.4 | 8.0±1.2 |
| Palmeira das Missões | 177 | June 18 | 179±16.0 | 516 | 30.4±5.3 | 8.8±1.5 |
| Santo Ângelo | 159 | June 23 | 162±15.0 | 581 | 28.0±5.0 | 10.0±1.8 |
| Erechim | 115 | July 9 | 114±12.8 | 488 | 26.0±6.4 | 11.1±2.7 |
| Ijuí | 110 | August 9 | 111±12.7 | 483 | 22.1±4.8 | 9.6±2.1 |
| Santa Rosa | 109 | August 2 | 112±12.3 | 498 | 19.3±3.8 | 8.6±1.7 |
| Cruz Alta | 101 | July 2 | 103±11.8 | 681 | 17.0±3.1 | 11.2±2.0 |
| Santa Cruz do Sul | 84 | July 12 | 88±10.6 | 251 | 14.4±2.7 | 4.1±0.8 |
| Bagé | 54 | August 25 | 52±8.2 | 275 | 9.6±2.2 | 5.1±1.2 |
| Cachoeira do Sul | 45 | August 23 | 42±7.3 | 208 | 6.7±1.6 | 3.3±0.8 |

SARS-CoV-2 transmission varied in time, with an earlier increase of daily infections in those regions with highest attack rates (Fig. S1A). As expected from the structure of our model, the estimated number of deaths per day closely matches the number of notified deaths (Fig. S1B), but there was substantial underreporting of SARS-CoV-2 infections, shown by the gap between estimated (blue line) and reported (brown bars) cases in Figures 1A and S1A. The effective reproduction number (*R*_*t*_; Fig. S1C) also varied in time and among regions. The longest time that any region sustained values of *R*_*t*_ lower than 1 was of nine weeks, in Passo Fundo and Pelotas (Fig. 1). In Capão da Canoa, Guaíba, and Lajeado, *R*_*t*_ stayed below 1 throughout eight consecutive weeks. These five periods of low *R*_*t*_ happened between early August and mid-October, after the first peak of notified COVID-19 deaths in RS.

**Figure 1.**
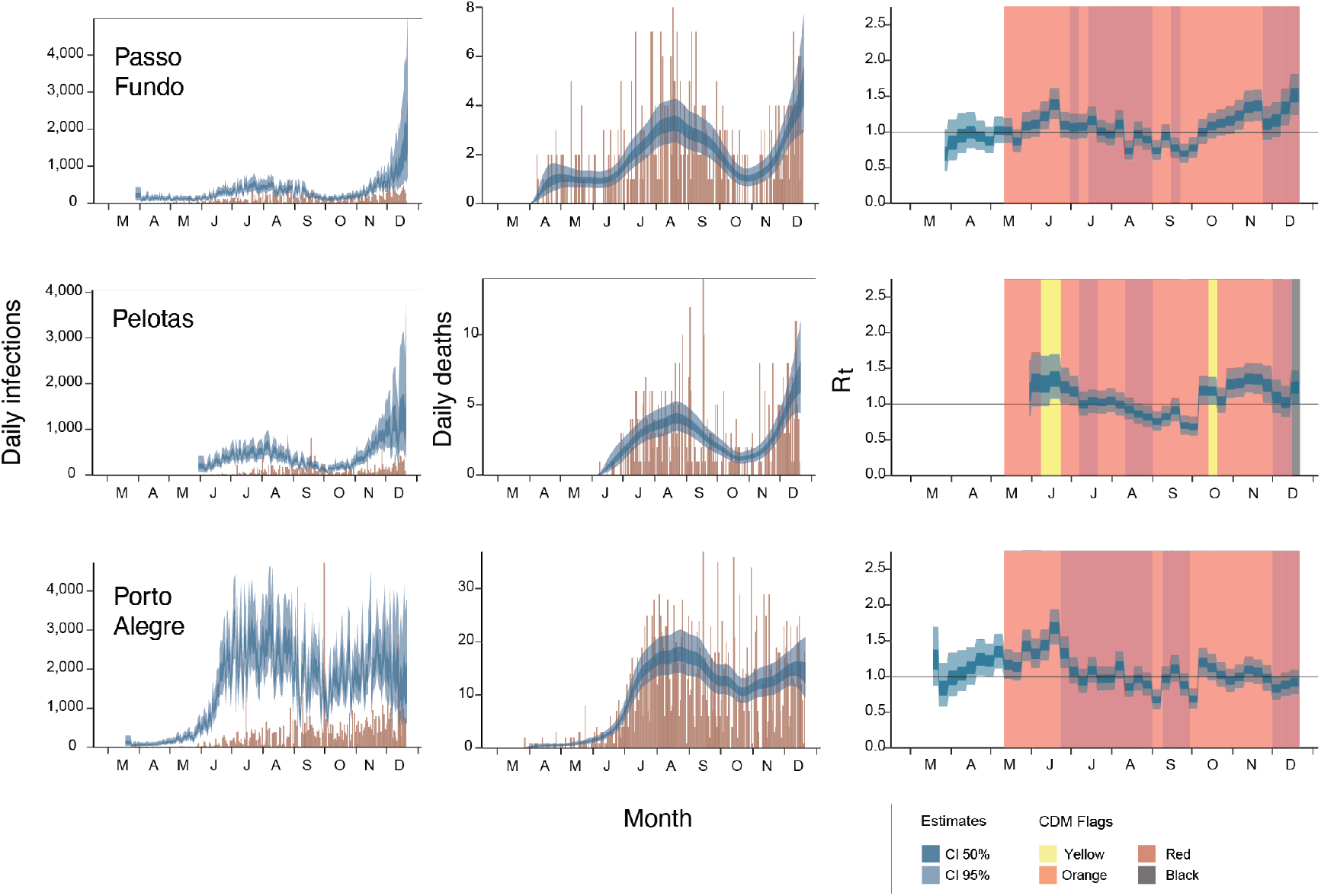
Temporal trajectories of SARS-CoV-2 daily infections (a), daily deaths due to COVID-19 (b) and weekly *R*_*t*_ (c) for the COVID region of Passo Fundo, Pelotas, and Porto Alegre throughout the study period. Panels A and B show notified numbers in brown and estimated numbers in blue (with light blue showing 95% and dark blue 50% credible intervals around the posterior mean). Likewise, the *R*_*t*_ values in panel C show, 95 and 50% credible intervals, respectively in light and dark blue. Vertical colored rectangles show the period of application for each CDM flag color. Weekly *R*_*t*_ values are a geometric mean of the daily estimates for the corresponding week. Supplementary Figure S1 shows equivalent plots for all 21 COVID regions of RS.

We found no evidence that changes in CDM flag color bore any influence on temporal variation of *R*_*t*_. The posterior distribution of *R*_*t*+1_/*R*_*t*_ when flags were upgraded to a more restrictive color in week *t*+1 was qualitatively the same as when flags were downgraded or unchanged (Fig. 2A). The same held true with a one-week lag between flag and *R*_*t*_ transitions, *i*.*e*. considering flag assignments for week *t*. The logistic regression slope parameter for the effect of *R*_*t*_ ratio on the probability of flag upgrade was negative with a one-week lag and positive with a lag of three weeks (Fig. 2B).

**Figure 2.**
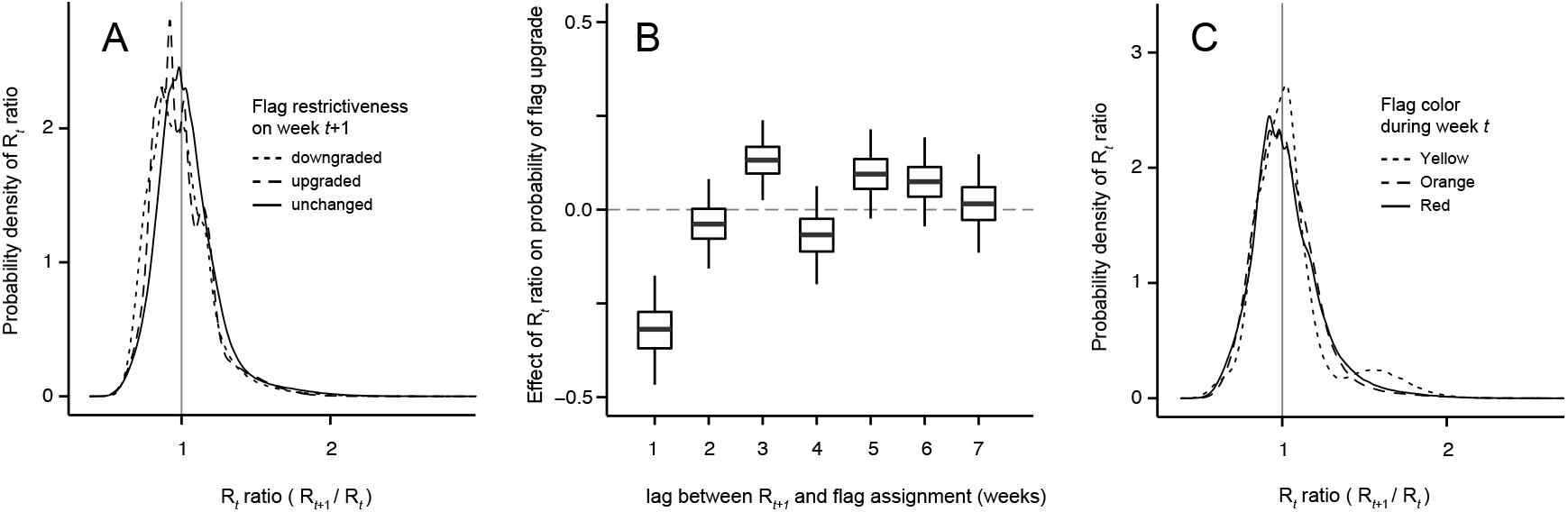
Relationship between *R*_t_ and CDM flag color, or lack thereof. Probability distribution of *R*_t_ ratio (measuring change from week *t* to *t*+1) is the same when flags are upgraded, downgraded, or stay the same in week *t*+1 (A). The probability of a flag upgrade increases with *R*_t_ ratio three weeks (but not one or two weeks) after *t*+1 (B). The probability distribution of *R*_t_ ratio does not change with flag color on week *t* (C).

Our data comprise 602 region-specific transitions between subsequent weeks, each with a corresponding flag color in the final week. The ranking from most to least frequently assigned color was orange, red, yellow and black (Table S3). Black flags were only assigned twice on two different regions, for one week each, with *R*_*t*_ increasing during the corresponding period in both cases. Figure 2C shows how the posterior distributions of *R*_*t*_ ratio derived for periods with yellow, orange, and red flag on week *t* are statistically undistinguishable from each other. Since red-flag periods provide the best basis for investigating effects of the most restrictive CDM interventions, we also explored the relationship between red-flag period duration (in weeks) and the corresponding change in *R*_*t*_ (Fig. 3). Only four out of eight periods of more than five weeks coincide with a substantial decrease. Of these, the three longest, with ten weeks each, in Canoas, Porto Alegre and Novo Hamburgo, were the most substantial. Twenty four out of thirty red-flag periods lasting between three and seven weeks showed either no evidence of alteration or an increase in *R*_*t*_.

**Figure 3.**
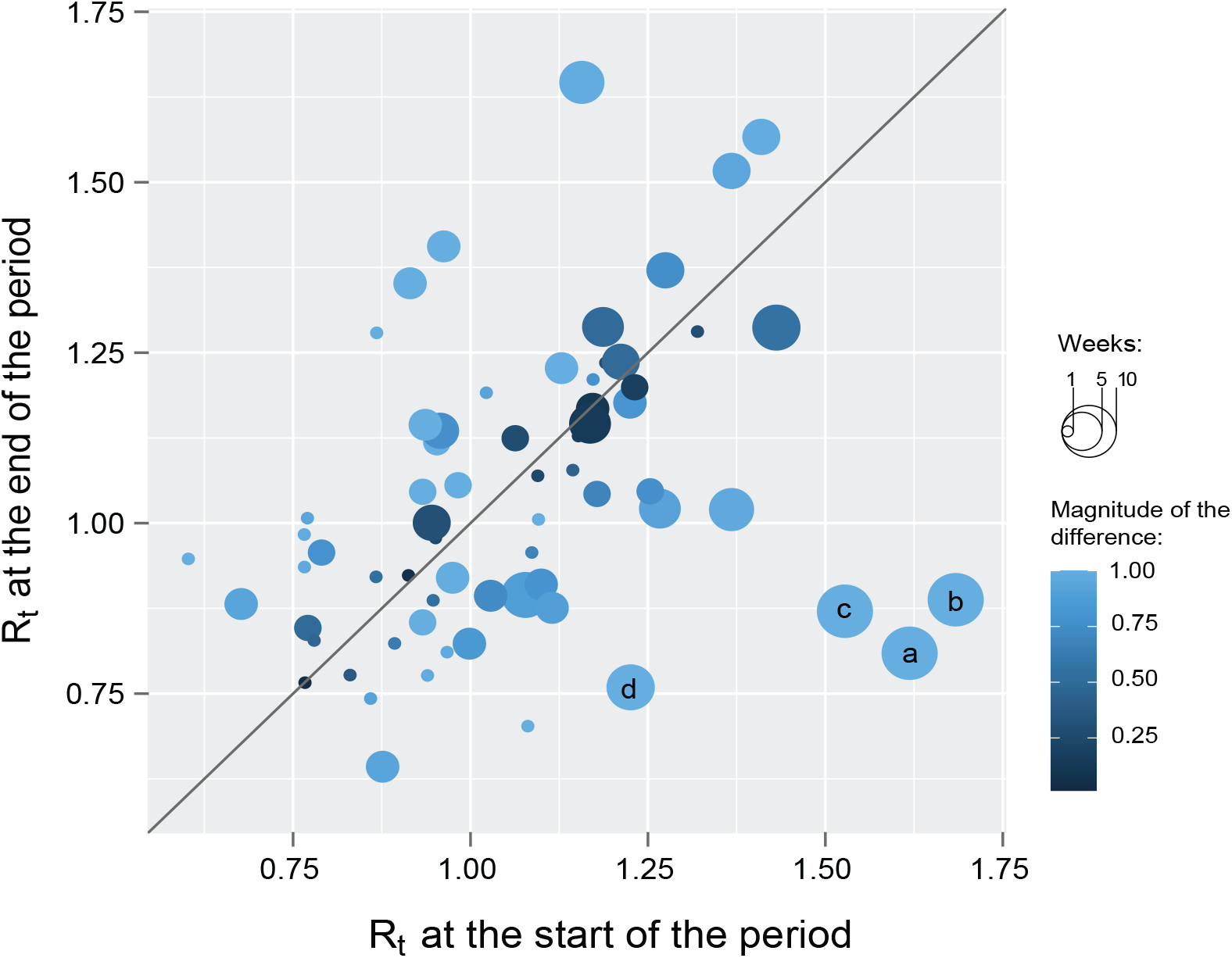
Relationship between the duration of continuous red-flag periods and change in *R*_*t*_ from the beginning to the end of the period, for all COVID regions. Circle sizes indicate period duration in weeks (Weeks) and their position gives the posterior mean of the *R*_*t*_ distributions identified by each of the axes. Circle colors indicate the proportion of the final *R*_*t*_ a posteriori distribution that is above (for *R*_*t*_ increases) or below (decreases) the diagonal line. Lighter colors indicate stronger evidence of *R*_*t*_ change. Letters *a, b, c* and *d* identify four periods with the strongest reductions in *R*_*t*_, from, respectively, the COVID regions of Canoas, Porto Alegre, Novo Hamburgo and Taquara.

We found no evidence whatsoever of a relationship between flag color and human mobility. Figure 4 shows a reduction in human mobility around the place categories of retails and recreation (Fig. 4A), parks (Fig. 4C), and public transport (Fig. 4D), in comparison with the reference period of January 03 to February 06, 2020. Concurrently, people spent more time in residential areas (Fig. 4F). The changes in mobility illustrated by Figure 4, however, show no relationship with flag color. This lack of relationship is supported by the linear mixed-effects model estimates of the effects of flag color on mobility (in all six place categories). The posterior distribution of these effects centers on zero and is statistically indistinguishable among place categories (Fig. S2).

**Figure 4.**
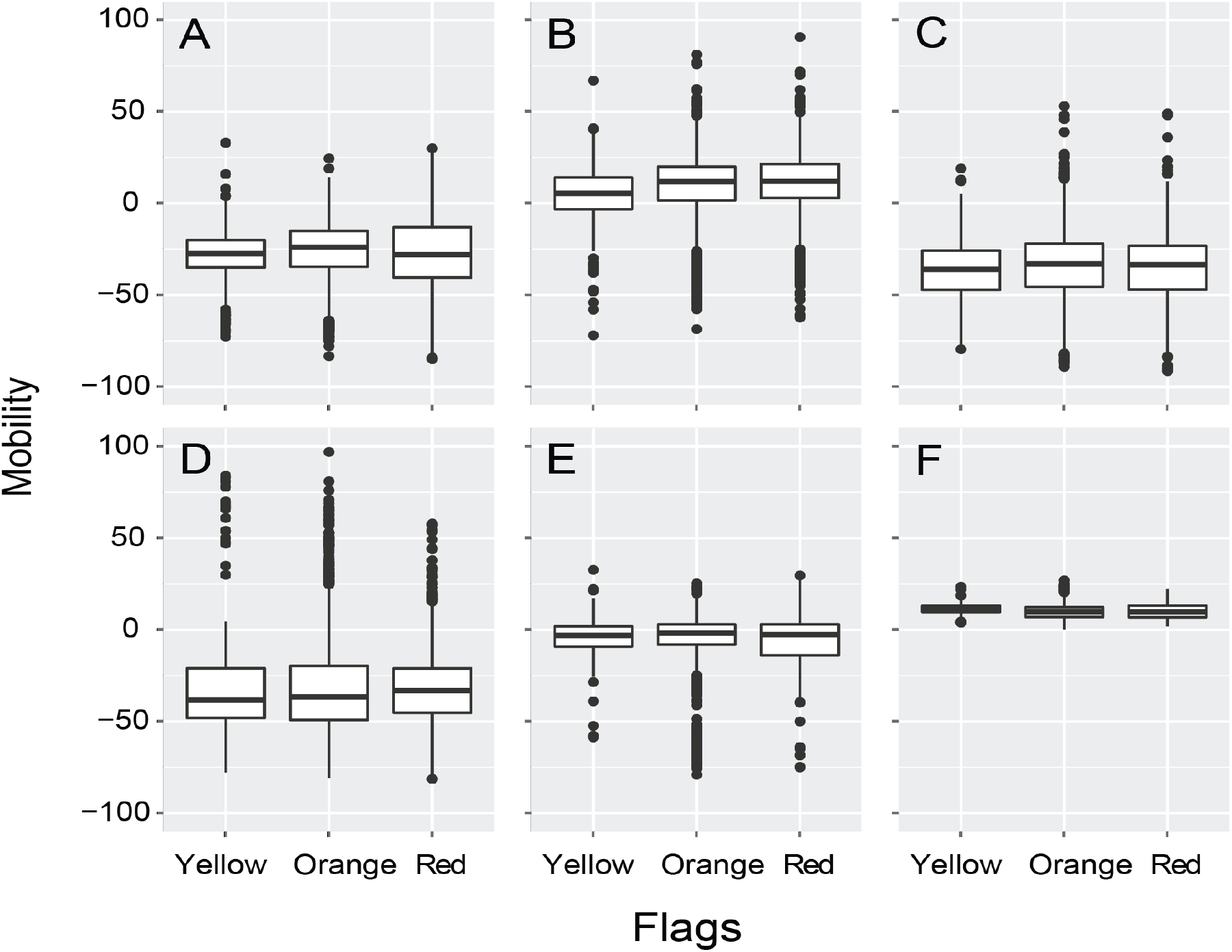
Boxplots of the six Google metrics of human mobility in the place categories of Retail and Recreation (a), Grocery and Pharmacy (b), Parks (c), Public Transport (d), Workplaces (e), and Residential areas (f). Each plot shows the distribution of daily metrics per COVID region across the three CDM flag colors that were most used throughout the study period. Metrics express proportional change from the reference period of January 3 - February 6, 2020. The metric of movement in residential areas quantifies proportional change in the average time that individuals spend at their residence; all the other metrics quantify proportional change in the number of visitors to a given place category.

## Discussion

We found extremely limited evidence that *R*_*t*_ responded to flag assignments, and some evidence that flag assignments reflected changes in *R*_*t*_, albeit with a three-week delay. The posterior distribution of *R*_*t*_ ratios were the same when flags were downgraded, unchanged or upgraded, both for the week of flag assignment and for the subsequent week. Likewise, *R*_*t*_ ratios were equally distributed during the week of assignment and the week after assignment of yellow, orange, and red flags. The only suggestion of *R*_*t*_ responsiveness to the CDM flags came from four extended periods of continuous red flag (8-10 weeks) in the Porto Alegre metropolitan area towards the middle of 2020. As for flag assignments reflecting *R*_*t*_, the probability of flag upgrade increased with increased values of *R*_*t*_ ratio, but only three weeks after the corresponding change in effective reproduction number. This lag broadly matches the sum of the periods from infection to onset of symptoms (∼5.1d) and from onset of symptoms to death (∼15.4d) reported in the literature.^20-22^ Without contact tracing nor testing of people without symptoms, the CDM was only able to assess increases in transmission when people sought help from the health services or died, which would explain the three-week delay in flag-assignment response. The fact that peak-*R*_*t*_ values must be followed by low *R*_*t*_ ratios, helps explain the negative relationship between ratio and flag upgrade observed at the shortest time lag. Flags were upgraded when the health system was being strained and *R*_*t*_ was already on a downslope.

Like any inference based on statistical modeling, our estimates of *R*_*t*_ rely on assumptions that must be pondered. Because we modeled infection based on deaths data, the infection fatality ratio (IFR) is an important element of our study. Sensitivity analyses revealed that IFR does influence the number of infections estimated by a model like ours^13^, but IFR is robust to changes in the contact matrix^12^. We derived one value of IFR for each COVID region while assuming region-specific age structures and a shared contact matrix. The resulting values were within the range of published IFR estimates for RS.^27^ Likewise, the assumed distributions of incubation period, time between onset of symptoms and death, and serial interval have been corroborated in the literature.^28^ Perhaps the riskiest assumption of our analysis is to hold *R*_*0*_ constant (within region) throughout the study period. The P.1 SARS-CoV-2 lineage that emerged in Manaus mid-November, 2020, was more transmissible than previously circulating lineages.^29^ With *R*_*t*_ estimated as a function of *R*_*0*_ and human mobility metrics, an *R*_*0*_ bias could conceal a relationship between *R*_*t*_ and flag colors—that is, if P.1 became sufficiently prevalent before the end of 2020. Lineage P.1 did account for more than 85% of sequences sampled at the largest RS hospital in February 2021,^30^ but, as of this writing, it was found on only 3 out of 263 RS sequences in the GSAID repository from the last two months of 2020^31^. We nonetheless re-fitted our model to data collected until October 26, before the earliest possible date of P.1 emergence in RS. The results, summarized in figure S3, were qualitatively similar to those reported here.

The three-week lag between *R*_*t*_ increase and associated flag upgrade suggests the CDM was detecting relevant changes in transmission, but doing so too late for a timely response. In fact, the strongest evidence for a relationship between *R*_*t*_ ratio and probability of flag upgrade is the negative relationship at one-week lag. That is, delayed assessment of an increase from three weeks prior often masked the evidence of recent reduction in *R*_*t*_. How could the lag be shortened? Certainly not with an approach reliant on deaths data, appropriate for a retrospective study. The challenges of estimating *R*_*t*_ in real time are such^32-33^ that some authors consider replacing it with epidemic growth rates,^34-36^ but renewal-equation models, like ours, can also be fit to case data.^37^ Such models do shorten the lag between assessment and transmission, but their usefulness depends on data quality. Ideally, models for estimating *R*_*t*_ (or epidemic growth rate, for that matter) should integrate real-time data from different sources, including different types of testing and environmental monitoring, as in recent variant prevalence studies.^38^

Apart from the four long periods of red flag that saw *R*_*t*_ decreasing to values below 1 in Porto Alegre, Novo Hamburgo, Canoas, and Taquara (Fig. 3), we found no relationship between the CDM flag system and the pace of disease transmission: 1) *R*_*t*_-ratio distributions were similar regardless of whether flags were upgraded, downgraded, or unchanged in weeks *t*+1 or *t*; 2) *R*_*t*_-ratio distributions were similar regardless of flag color on week *t*; and 3) duration of red flag periods (apart from the four cited exceptions) bore no relationship with the magnitude or direction of *R*_*t*_ change, thus suggesting that flag colors did not affect disease spread neither in the short nor in the medium term. This must not be interpreted as evidence that NPIs do not affect transmission. Indeed, visual examination (Fig. 4) and a linear mixed-effects model (Fig. S2) of the relationship between flag colors and human mobility strengthen the notion that people generally ignored the flags. Whatever changes took place in human mobility with respect to the reference period of January 2020, which were real,^2,39^ happened irrespective of the CDM. From the response point of view, the framework was insufficient to stem the transmission of SARS-CoV-2 because, since it failed to impress human behavior, the intended NPIs did not materialize.

The long red-flag periods in Canoas, Porto Alegre, and Novo Hamburgo, lasted from June 23^rd^ to August 31^st^. Their ending illustrates how the CDM struggled to contain dissemination of SARS-CoV-2 in RS. Between July 15^th^ and August 31^st^, the government enforced a mandate for applying black and red flags during at least two weeks once they had been assigned to a region. This mandate was withdrawn in the first week of September, when all three regions had *R*_*t*_ estimates with 95% credible intervals completely below 1. In the same week, all three regions reverted to orange flag, occasionally returning to red but never for more than three weeks in a row. By mid-October, they all had estimates of *R*_*t*_ > 1. Assessment-response frameworks can be powerful tools for responding to temporal change in healthcare systems,^40^ but given the challenges of estimating *R*_*t*_ in near-real time, especially during resurgence periods,^33^ we concurr with the notion^33^ that assessment is more useful for deciding when to stop than when to start implementation of NPIs. When the protection of lives is seen as inseparable from the protection of livelihoods,^41^ one may expect sufficient societal coordination to prevent uncontroled growth of an epidemic with NPIs implemented before detection of dangerously high transmission. When prevention fails, assessment of transmission can and should be put in place for deciding when to interrupt NPI implementation.

## Data Availability

All data produced are available online at https://osf.io/gxfq5/

## Acknowledgments

We thank Thomas Mellan and the Imperial College COVID-19 Response Team for sharing code and providing feedback throughout the process of adapting and fitting the epidemiological model that is at the core of this paper. Fernando Abad-Franch and Glauco Machado offered insightful comments on earlier versions of the manuscript. This study was funded by an MSc grant (#88887.502993/2020-00) from the Brazilian Coordination for the Improvement of Higher Education Personnel (CAPES); as well as by PhD (#400559/2023-4), Productivity (#312519/2021-4), and research (#403651/2022-0) grants from the Brazilian National Council for Scientific and Technological Development (CNPq).

## Notes

### Competing Interest Statement

The authors have declared no competing interest.

### Author Declarations

This research was submitted to Plataforma Brasil, and approved by the Ethics Committee 5337 - Santa Casa da Misericordia de Pelotas under the Certificate of Presentation of Ethical Appreciation (CAAE) 34335920.1.0000.5337 on October 9, 2020. This research was conducted without access to any individual information.

