## Supplemental Materials for "SARS-CoV-2 Spread Under the Controlled-Distancing Model of Rio Grande do Sul, Brazil"

### Supplementary Methods

The epidemiological model employed in our analysis follows a hierarchical Bayesian approach that estimates the effective reproduction number ($R_t$) conditioned on observed deaths. The model includes three components: 1) a deaths component; 2) an infections component; and 3) a component that estimates the effective reproduction number as a function of human mobility. All three parts incorporate temporal variation in parameter estimates. Components are interlinked, so that estimation of all parameters happens simultaneously, as the whole model is fit to data. Below, we offer a detailed description of each model component and the relationships among model parameters. Our epidemiological model is an adaptation of previous work developed by the Imperial College COVID-19 Response Team and collaborators, who established a latent renewal equation approach that combines the three parts listed above.[1] That approach has been incremented to include the effect of mobility metrics on $R_t$ estimates,[2] as well as a weekly autoregressive process for estimating $R_t$,[3] which improves flexibility in adjusting estimates to data volume. The stan code for the epidemiological model is embedded in *Supplementary Code 1* (below) and the *Code_1.R* file. Code for exploring the relationship between Rt estimates and the CDM flags is given in *Supplementary Code 2* (below) and the *Code_2.R* file. We also present a description of the linear mixed-effects model to explore the relationship between CDM flag colors and google mobility metrics from Rio Grande do Sul (RS); this is available in *Supplementary Code 3* and the *Code_3.R* file. Each R code comes with a corresponding RData file with the necessary data.

#### Data

Our study of SARS-CoV-2 transmission in the 21 RS COVID regions combines epidemiological, demographic, mobility, and control data collected between 15 March and 21 December 2020. Epidemiological data on daily reported infections and deaths were obtained from the *Painel Coronavírus do Ministério da Saúde*.[4] Demographic data, in the form of projected human population age structure per municipality, came from the digital archive of the now extinct state government agency *Fundação de Economia e Estatística do Rio Grande do Sul*.[5] Municipality-level mobility metrics came from the website of Google's COVID-19 Community Mobility Reports.[6] Google provides metrics of change in daily mobility with reference to January 2020 for six place categories: Residential, Transit stations, Parks, Grocery & pharmacy, Retail & recreation, and Workplaces. We computed regional-level mobility metrics as averages of the respective municipality-level values weighted by each municipality's population size. Weekly, region-specific control data, expressed by CDM flag colors, were obtained from the official site of the RS government.[7]

#### Deaths Component of the Epidemiological Model

We modelled $D_{i,t}$, the observed number of deaths in COVID region *i* and day *t*, as a negative binomial distribution, as given by Flaxman *et al.*[1] and Mellan *et al.*:[2]

$$D_{i,t} \sim \text{Negative Binomial}\left(d_{i,t}, d_{i,t} + \frac{{d_{i,t}}^2}{\psi}\right),$$

where $d_{it}$ is the expected number of deaths in COVID region *i* and day *t*, and $\psi$ is an overdispersion parameter that follows a (positive) half-Normal distribution:

$$\psi \sim \text{Normal}^{\,+}(0{,}5).$$

The sum $d_{i,t} + \frac{{d_{i,t}}^2}{\psi}$ gives the variance of the $D_{it}$ distribution, used in the alternative parameterization of the negative binomial by the *neg_binomial_2* stan function.

The relationship between the numbers of deaths and the number of infection cases is given by a COVID-19 infection fatality rate ($IFR$, the probability of death given infection) and a set of epidemiological distributions: 1) a distribution of times from the onset of symptoms to death; 2) a distribution of times from infection to onset of symptoms; and 3) a distribution for the serial interval, the average time between a host's infection and its transmission of the pathogen to another host. We will detail the first two distributions here and the last one in the infection cases section.

Region-specific $IFR$ values were computed before fitting the model to data, following the procedure outlined by Flaxman et al. (see *Supplementary Code 4* and *Code_4.R* file).[1] This started with an estimate of $IFR$ obtained by Verity et al.[8] based on a sample of 44 672 laboratory-confirmed infection cases from China, observed between Jan 1 and Feb 11, 2020. This information was combined with infection prevalence data from 689 non-chinese Wuhan residents repatriated on six flights between Jan 30 and Feb 1, 2020. We took the mean of the resulting $IFR$ estimate and adjusted it according to region-specific human population age structure from our study area.[5] We also took into account a matrix of contact frequencies among people in different age classes from Peru—the closest geographical match to our study area,[9] and a basic reproduction number.[10] In the end, the procedure gave us a vector of region-specific $IFR$ values that are shown on *Supplementary Table S4*.

The time from onset of symptoms to death is given as a Gamma distribution (implemented with the alternative parameterization of R's *rgammaAlt* function) with mean 15.4 days[11] and coefficient of variation 0.45 [2]. The second distribution that we list above, for times of infection to onset of symptoms—the incubation period—is also Gamma, but with mean 5.1 days (an average of values reported by[12,13]) and coefficient of variation 0.86.[2] These two distributions jointly, see inside the parentheses below, express the distribution of times from infection to death in COVID region *i* as:

$$\pi_i \sim IFR_i\big(Gamma(5.1{,}0.86) + Gamma(15.4{,}0.45)\big),$$

where $IFR_i$ is the infection fatality rate for COVID region *i*.[1]

Finally, the expected number of deaths $d_{i,t}$, on COVID region *i* and day $t$, is given by the sum of products between a) the number of new infections on day $\tau$ before $t$, $c_{i,\tau}$, and b) the probability of dying from an infection in $t - \tau$ days:[1]

$$d_{i,t} = \sum_{\tau=0}^{t-1} c_{i\tau}\, \pi_{i,t-\tau}.$$

Here, the time-indexed probability of dying from an infection in region *i* corresponds to a discretization of $\pi_i$,[1] above, given by:

$$\pi_{i,t} = \int_{\tau=t-0.5}^{t+0.5} \pi_i\,(\tau) d\tau, \text{ for } t = 2, 3, \ldots,$$

And

$$\pi_{i,1} = \int_{\tau=0}^{1.5} \pi_i\,(\tau) d\tau.$$

**Infection Cases Component of the Epidemiological Model**

The infection component of our model is centered on the distribution of the average time from a host's infection to his transmitting the virus to another host, also known as the serial interval distribution. Still following Flaxman et al.[1] we model this distribution as gamma with a mean of 6.5 days and coefficient of variation of 0.62:

$$g \sim \text{Gamma}(6.5{,}0.62).$$

Thus, the number of infections $c_{it}$ in region *i* and day *t*, which appeared in the previous section, is described as a renewal process by the discrete convolution function:[1]

$$c_{it} = R_{it} \sum_{\tau=0}^{t-1} c_{i\tau}\, g_{t-\tau}.$$

The continuous distribution of times to transmission $g$ is being discretized into a per-day probability of transmission according to:

$$g_t = \int_{\tau=t-0.5}^{t+0.5} g(\tau)d\tau \text{ for } t = 2, 3, \ldots,$$

and

$$g_1 = \int_{\tau=0}^{1.5} g(\tau)d\tau.$$

The initial number of infections from which we started the analysis was given by

$$c_{1i}, \ldots, c_{6i} \sim \text{Exponential}(1/\tau),$$

where $\tau \sim \text{Exponential}(0.03)$.[2] In short and in words, the number of infections on a given day depends on the product of how many infections there were on previous days by the time it takes to transmit them, scaled by the region's effective reproduction number.

**Effective Reproduction Number**

The effective reproduction number of region *i* ($R_{it}$) is derived from its basic reproduction number ($R_{i0}$) adjusted by the impact of non-pharmaceutical interventions and a temporal autoregressive process (adapted from Flaxman et al.,[1] Mellan et al.[2] and Unwin et al.[3]):

$$R_{it} = R_{i0} 2\lambda^{-1} \left( \left( -\sum_{k=1}^{4} (\alpha_k + \beta_{ik})\, I_{ikt} \right) - \epsilon_{iw_i(t)} \right).$$

In this equation, $\lambda^{-1}$ denotes the logistic function, with coefficients $\alpha_k$ and $\beta_{ki}$ representing, respectively, the effect of mobility indicator *k* shared among all COVID regions and the effect of mobility indicator *k* specific to each COVID region *i*. $I_{ikt}$ is the value of the *k*'th mobility indicator for region *i* on day *t*. $\epsilon_{iw_i(t)}$ represents weekly autoregressive variation in region *i* that is not explained by variation in the mobility metrics.

Our prior distribution for $R_{i0}$ is normal with a mean of 3.28 as found by Liu et al.[10] and a standard deviation of 0.5. In earlier applications of this model, the standard deviation for this prior was given by the absolute value of a normally-distributed κ with mean 0 and standard deviation 0.5.[2]

That formulation of the prior presented convergence problems in our analysis, so we decided to simplify the prior for $R_{i,0}$ using the fixed value of 0.5 for its standard deviation. The mobility indicator, $I_{ikt}$, measures the impact of non-pharmaceutical interventions and shows the daily reduction or increase in mobility with respect to reference values of Jan 3 to Feb 6, 2020. As the k index changes from 1 to 4, the mobility indicator gives information about residential areas (*k* = 1), transit stations (*k* = 2), parks (*k* = 3), and the average of groceries & pharmacies, retail & recreational areas, and workplaces (*k* = 4). Mobility data were obtained from COVID-19 Community Google Mobility Reports, which compile indicators at the municipality level. In order to obtain daily regional values, we averaged the municipalities belonging to each region, weighted by their human population in 2017, as given by *Fundação de Economia e Estatística*.[5] We used priors for shared and region-specific effects adapted from Mellan et al.[2] as:

$$\alpha_k \sim \text{Normal}(0, 0.5)$$
$$\beta_{ki} \sim \text{Normal}(0, 0.125).$$

The mean and standard deviation of $\alpha_k$ are the same as used by Mellan et al..[2] The standard deviation (sigma) of $\beta_{ki}$ used by Mellan et al.[2] was a normally-distributed γ value, with mean 0 and standard deviation 0.5. We replaced this normal distribution of sigma with the fixed value of 0.125 to obtain convergence.

The last term of the logit-linear expression for $R_{it}$, $\epsilon_{iw_i(t)}$, is given on a weekly scale, so that $w_i(t)$ represents the week in region *i* that corresponds to day *t*. The transformation from daily to weekly temporal scale is given by $[(t - t_i^{start})/7] + 1$, where $t_i^{start}$ is the first day of seeding of infections on COVID region *i*. This effect represents an autoregressive process of order two,[3] because it draws on values from the latest two times in the weekly time series ($\epsilon_{w-1}$ and $\epsilon_{w-2}$), for $i \geq 2$, as

$$\epsilon_{wi} \sim \text{Normal}(\rho_1\epsilon_{w-1i} + \rho_2\epsilon_{w-2i}, {\sigma_w}^*)$$

in which the priors are given by

$$\rho_1 \sim \text{Normal}(0.8, 0.05),$$
$$\rho_2 \sim \text{Normal}(0.1, 0.05),$$
$$\text{and } \sigma_w \sim \text{Normal}^+(0,0.2).$$

The value of ${\sigma_w}^*$ is obtained by the product $\sigma_w\sqrt{1-\rho_1^2-\rho_2^2-2\rho_1^2\rho_2/(1-\rho_2)}$. The autoregressive process for *i* = 1 is given by

$$\epsilon_{1i} \sim \text{Normal}(0, {\sigma_w}^*)$$

starting 30 days before a total of 10 cumulative deaths has been observed in COVID region *i*.

We estimated parameters for the 21 COVID regions in a single hierarchical model fitted in Stan.[14] We ran 4 chains for 4000 iterations with 2000 iterations of warmup and a thinning factor of 1, to obtain 4000 posterior samples. Posterior convergence was assessed using the R-hat statistic and by diagnosing divergent transitions of the Hamiltonian Monte Carlo sampler. Data management and processing results was done in R.[15]

### Linear Mixed-Effects Model of Mobility and Flag Color

We explored effects of the CDM flag colors on mobility with a linear mixed model:[16]

$$y_{ti} = \alpha_{ti} + \beta_i + \gamma_t + \varepsilon_{ti},$$

where $y_{ti}$ represents the mobility indicator at time *t* from COVID region *i*, $\alpha_{ti}$ represents the flag's effect at time *t* from COVID region *i*, $\beta_i$ is the effect from COVID region *i*, and $\gamma_t$ is the temporal effect of adhesion to the system at time *t*. The flag's effect was assumed as fixed effect, with prior given by

$$\alpha_{ti} \sim \text{Normal}(0, 0.001),$$

while the region and adhesion effects were modeled as random effects, given by

$$\beta_i \sim \text{Normal}(\mu_\beta, \sigma_\beta),$$
$$\gamma_i \sim \text{Normal}(\mu_\gamma, \sigma_\gamma).$$

In the random effect specification, $\mu_\beta$ and $\mu_\gamma$ follow a Normal distribution with mean 0 and standard deviation 0.001, while $\sigma_\beta$ and $\sigma_\gamma$ are given by $\sigma = 1/\tau^2$, in which $\tau$ follows a uniform distribution between 0 and 100.

**Table S1.** Description of the eleven metrics of the Controlled-Distancing Model.

| Set | Regional Level | Weight | Metric |
|---|---|---|---|
| Virus Propagation | 21 Regions | 0.375 | Ratio of the number of new COVID-19 confirmed cases[1] during the last seven days over the number of new COVID-19 confirmed cases during the preceding week (eighth to fourteenth day before the reference day) |
| | 7 Macroregions | 0.375 | Ratio of the number of the ICU SARS inpatients at the reference day over the number of the ICU SARS inpatients seventh days before the reference day |
| | 7 Macroregions | 0.375 | Ratio of the number of COVID-19 patients in clinical beds on the reference day over the number of COVID-19 patients in clinical beds on the seventh day before the reference day |
| | 7 Macroregions | 0.375 | Ratio of the number of COVID-19 patients in ICU on the reference day over the number of COVID-19 patients in ICU on the seventh day before the reference day |
| | 21 Region | 1 | Ratio of the number of COVID-19 active cases[2] over the number of recoveries from COVID-19 during the last fifty days[3] |
| | 21 Region | 1.25 | Number of confirmed COVID-19 cases[4] per 100,000 inhabitants during the seven days prior to the reference day |
| | 21 Region | 1.25 | Number of COVID-19-attributed deaths during the seven days prior to the reference day, per 100,000 inhabitants[5] |
| Healthcare service capacity | 7 Macroregions | 1.25 | Number of ICU beds available for COVID-19 patients on the reference day per 100,000 elderly citizens[6] |
| | State | 1.25 | Number of ICU beds available for COVID-19 patients on the reference day[7] |
| | 7 Macroregions | 1.25 | Ratio of the number of ICU beds available for COVID-19 patients at the reference day over the number of ICU beds available for COVID-19 patients on the seventh day prior to the reference day[8] |
| | State | 1.25 | Ratio of the number of ICU beds available for COVID-19 patients on the reference day over the number of ICU beds to available COVID-19 patients on the seventh day prior to the reference day[9] |

[1] Number of COVID-19 confirmed cases (the ratio's numerator) was replaced by the number of COVID-19-positive hospitalizations on 25 May, 2020. Later, on 15 June, 2020, the numerator itself was replaced by a ratio, this time of the number of active cases during the seven days prior to the reference day over the number of recovered cases during the same period.
[2] Cases with positive results from tests performed with samples collected within the last fourteen days before the reference day.
[3] On 15 June, 2020, this metric was replaced by the total number of active cases in the region during the last week divided by the number of recoveries in the region throughout the fifty days prior to the reference day.
[4] On 25 May, 2020, number of confirmed COVID-19 cases was replaced by number of COVID-19-positive hospitalizations.
[5] On 15 June, 2020, replaced by projected number of deaths in the region for one week, per 100,000 inhabitants.
[6] On 15 June, 2020, this metric was replaced by the number of ICU beds available in the microregion for treating COVID-19 patients divided by the number of ICU beds occupied by COVID-19 patients in the macroregion on the latest measurement day.
[7] Replaced on 15 June, 2020, by the number of ICU beds available in the state for treating COVID-19 patients, divided by the number of ICU beds occupied in the state by COVID-19 patients.
[8] On 15 June, 2020, replaced by the ratio of ICU beds available for treating COVID-19 in the macroregion on the reference day over the number of ICU beds available for treating COVID-19 in the macroregion seven days prior to the reference day.
[9] Replaced, on 15 June, 2020, by the ratio of the number of ICU beds available for treating COVID-19 in the state on the reference day over the number of ICU beds available for treating COVID-19 in the state seven days prior to the reference day.

**Table S2.** Main decrees issued by the state government of Rio Grande do Sul to regulate the state's response to the COVID-19 pandemic, including implementation of the controlled-distancing model, with their number, date, and description.

| State Decree # | Date | Description |
|---|---|---|
| 55128 | March 19, 2020 | Declaration of the state of public calamity and implementation of the first non-pharmaceutical interventions. |
| 55240 | May 10, 2020 | Establishment of the controlled-distancing model (CDM). |
| 55320 | June 20, 2020 | Implementation of block mechanism to prevent short-term downgrading of CDM-attributed red or black flags. |
| 55435 | August 11, 2020 | Establishment of sanitary protocol co-management between state and municipal governments. |
| 55460 | August 31, 2020 | Revocation of the block mechanism implemented by decree # 55320, above. |
| 55882 | May 15, 2021 | Replacement of the CDM by a new, more-flexible framework for controlling COVID-19 in the state. |

**Table S3.** Number of weeks per flag color and COVID region, from the beginning until the 32nd week of the CDM implementation. Letters 'P' and 'D' show, respectively, the –'preliminary' and 'definitive' flags. Every week, preliminary flags were announced on Friday by the government, pending appeal by municipalities which often negotiated a different flag color for definitive implementation in their regions by the following Monday.

| COVID region | Flag | | | | | | | |
|---|---|---|---|---|---|---|---|---|
| | Yellow | | Orange | | Red | | Black | |
| | P | D | P | D | P | D | P | D |
| Bagé | 10 | 10 | 16 | 19 | 5 | 2 | 1 | 1 |
| Cachoeira do Sul | 7 | 7 | 20 | 22 | 5 | 3 | 0 | 0 |
| Canoas | 0 | 0 | 15 | 16 | 17 | 16 | 0 | 0 |
| Capão da Canoa | 1 | 1 | 16 | 16 | 15 | 15 | 0 | 0 |
| Caxias do Sul | 0 | 0 | 18 | 25 | 14 | 7 | 0 | 0 |
| Cruz Alta | 0 | 0 | 20 | 23 | 12 | 9 | 0 | 0 |
| Erechim | 1 | 1 | 19 | 24 | 12 | 7 | 0 | 0 |
| Guaíba | 0 | 0 | 14 | 22 | 18 | 10 | 0 | 0 |
| Ijuí | 4 | 4 | 18 | 20 | 10 | 8 | 0 | 0 |
| Lajeado | 0 | 0 | 23 | 26 | 9 | 6 | 0 | 0 |
| Novo Hamburgo | 0 | 0 | 14 | 15 | 18 | 17 | 0 | 0 |
| Palmeira das Missões | 2 | 2 | 13 | 17 | 17 | 13 | 0 | 0 |
| Passo Fundo | 0 | 0 | 16 | 19 | 16 | 13 | 0 | 0 |
| Pelotas | 3 | 3 | 19 | 21 | 9 | 7 | 1 | 1 |
| Porto Alegre | 0 | 0 | 13 | 16 | 19 | 16 | 0 | 0 |
| Santa Cruz do Sul | 3 | 3 | 21 | 25 | 8 | 4 | 0 | 0 |
| Santa Maria | 0 | 0 | 24 | 27 | 8 | 5 | 0 | 0 |
| Santa Rosa | 5 | 5 | 16 | 20 | 11 | 7 | 0 | 0 |
| Santo Ângelo | 0 | 0 | 16 | 21 | 16 | 11 | 0 | 0 |
| Taquara | 8 | 8 | 12 | 15 | 12 | 9 | 0 | 0 |
| Uruguaiana | 2 | 2 | 22 | 24 | 8 | 6 | 0 | 0 |
| **Total** | **46** | **46** | **365** | **433** | **259** | **191** | **2** | **2** |

**Table S4.** Infection fatality rates (IFR) and proportions of population above given ages for all the Rio Grande do Sul COVID regions.

| COVID region | IFR | >60 years | >65 years | >70 years | >75 years |
|---|---|---|---|---|---|
| Novo Hamburgo | 0.56% | 12.40% | 8.10% | 5.00% | 2.90% |
| Taquara | 0.59% | 13.00% | 8.80% | 5.50% | 3.20% |
| Canoas | 0.60% | 13.70% | 9.00% | 5.60% | 3.30% |
| Caxias do Sul | 0.66% | 14.40% | 9.80% | 6.30% | 3.80% |
| Guaíba | 0.66% | 15.00% | 10.20% | 6.50% | 3.80% |
| Porto Alegre | 0.70% | 15.60% | 10.60% | 6.90% | 4.30% |
| Capão da Canoa | 0.72% | 17.20% | 11.70% | 7.40% | 4.30% |
| Passo Fundo | 0.72% | 16.50% | 11.30% | 7.30% | 4.40% |
| Uruguaiana | 0.73% | 16.40% | 11.60% | 7.60% | 4.60% |
| Santa Cruz do Sul | 0.74% | 16.80% | 11.50% | 7.50% | 4.60% |
| Bagé | 0.74% | 16.60% | 11.70% | 7.70% | 4.90% |
| Cruz Alta | 0.76% | 17.20% | 12.00% | 7.90% | 4.80% |
| Lajeado | 0.77% | 17.40% | 12.10% | 8.00% | 4.90% |
| Pelotas | 0.77% | 17.60% | 12.30% | 7.90% | 4.90% |
| Ijuí | 0.78% | 17.90% | 12.50% | 8.20% | 5.00% |
| Palmeira das Missões | 0.79% | 18.30% | 12.60% | 8.20% | 5.10% |
| Santo Angelo | 0.80% | 18.40% | 12.80% | 8.40% | 5.10% |
| Santa Maria | 0.81% | 18.40% | 12.90% | 8.60% | 5.30% |
| Cachoeira do Sul | 0.82% | 18.90% | 13.20% | 8.70% | 5.30% |
| Erechim | 0.83% | 18.60% | 13.10% | 8.70% | 5.50% |
| Santa Rosa | 0.84% | 19.70% | 13.70% | 8.80% | 5.30% |

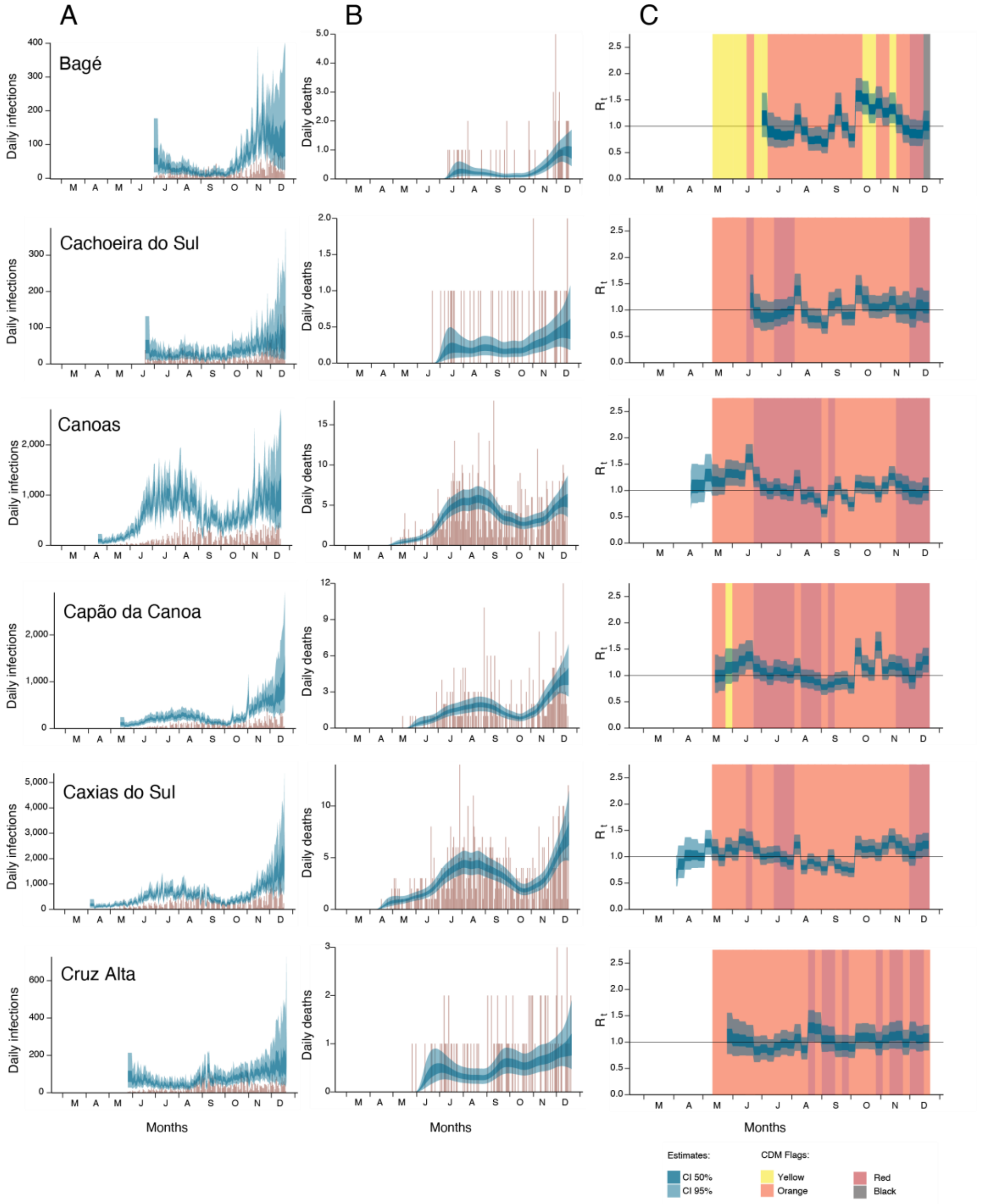

**Figure S1.** Temporal trajectories of SARS-CoV-2 daily infections **(a)**, daily deaths due to COVID-19 **(b)** and weekly $R_t$ **(c)** for each COVID region throughout the study period. Panels A and B show notified numbers in brown and estimated numbers in blue (with light blue showing 95% and dark blue 50% credible intervals around the posterior mean). Likewise, the $R_t$ values in panel C show, 95 and 50% credible intervals, respectively in light and dark blue. Vertical colored bars show the period of application for each CDM flag color. Weekly $R_t$ values are a geometric mean of the daily estimates for the corresponding week.

**Fig. S1.** (Contd.)

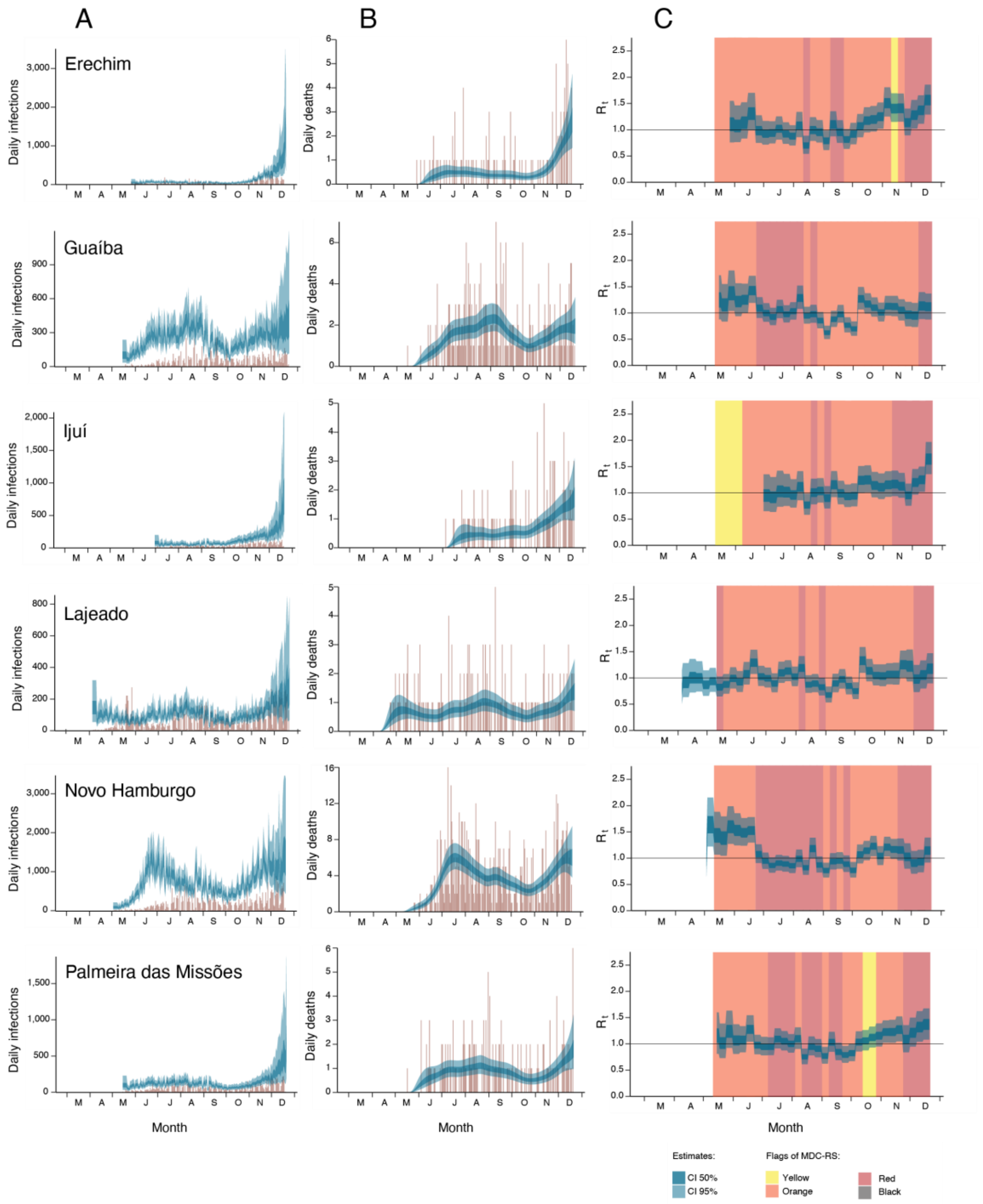

A
B
C
Erechim
Guaíba
Ijuí
Lajeado
Novo Hamburgo
Palmeira das Missões
Daily infections
Daily deaths
$R_t$
Month
Estimates:
CI 50%
CI 95%
Flags of MDC-RS:
Yellow
Orange
Red
Black

**Fig. S1.** (Contd.)

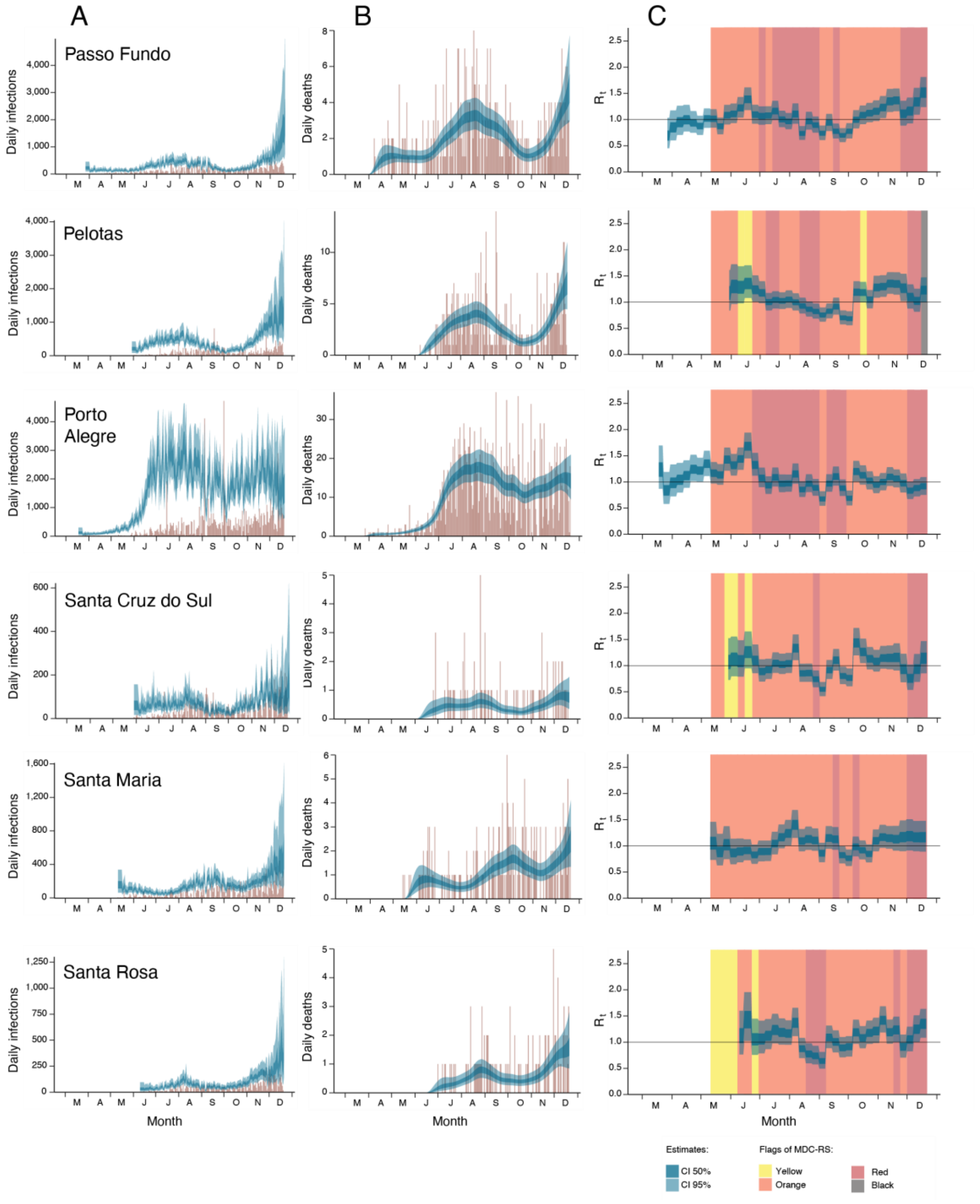
A
B
C
Passo Fundo
Pelotas
Porto Alegre
Santa Cruz do Sul
Santa Maria
Santa Rosa
Daily infections
Daily deaths
$R_t$
Month
Estimates:
CI 50%
CI 95%
Flags of MDC-RS:
Yellow
Orange
Red
Black

**Fig. S1.** (Contd.)

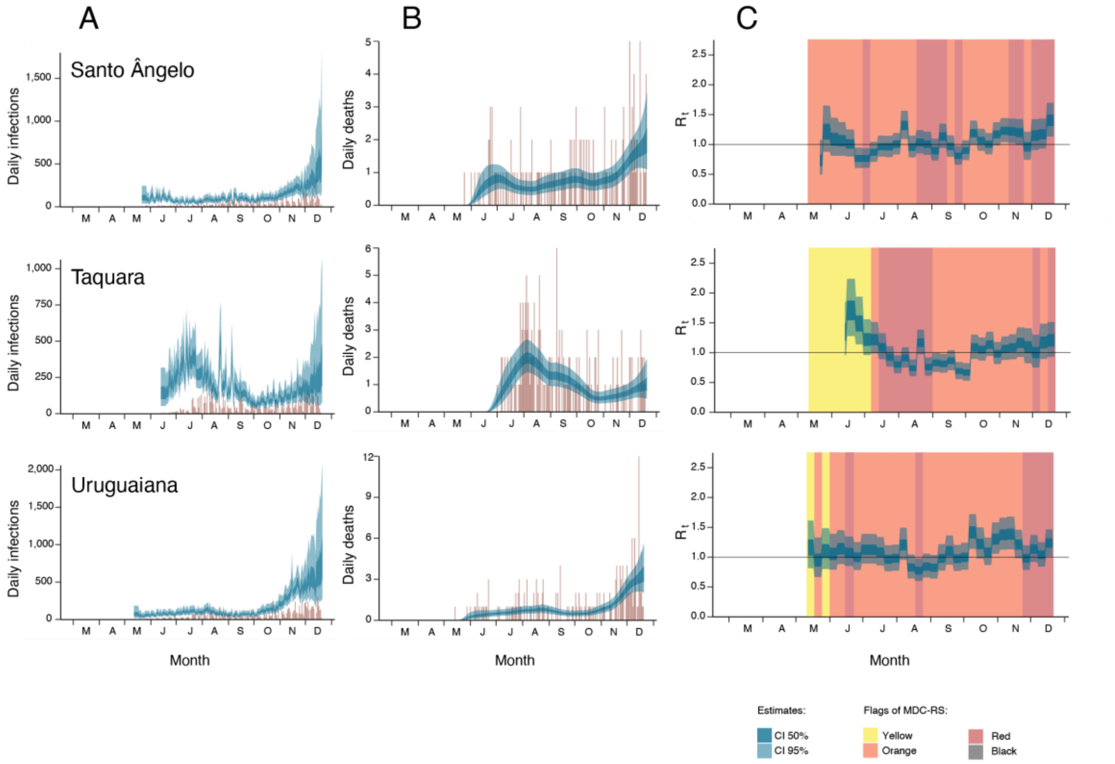

A
B
C
Santo Ângelo
Taquara
Uruguaiana
Daily infections
Daily deaths
$R_t$
Month
Estimates:
CI 50%
CI 95%
Flags of MDC-RS:
Yellow
Orange
Red
Black

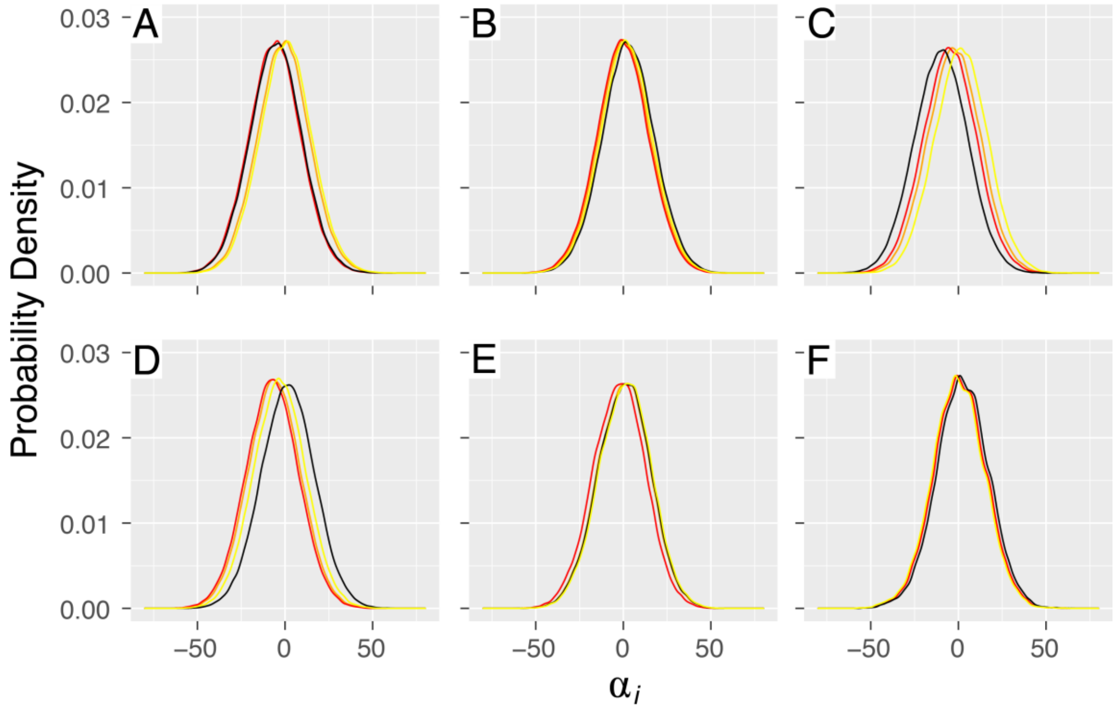

**Figure S2.** Posterior probability distribution of different flag effects on the six Google metrics of human mobility, according to the linear mixed-effects model. Effects are given by parameter $\alpha_i$, where $i$ stand for yellow, orange, red, and black flag colors, also given by the line color in the plot. Mobility metrics are taken in the six place categories of Retail and Recreation (**a**), Grocery and Pharmacy (**b**), Parks (**c**), Public Transport (**d**), Workplaces (**e**), and Residential areas (**f**). Negative values of $\alpha_i$ would indicate a reduction and positive values an increase in human mobility. Effective restrictive measures should result in reduced mobility everywhere but in residential areas (**f**).

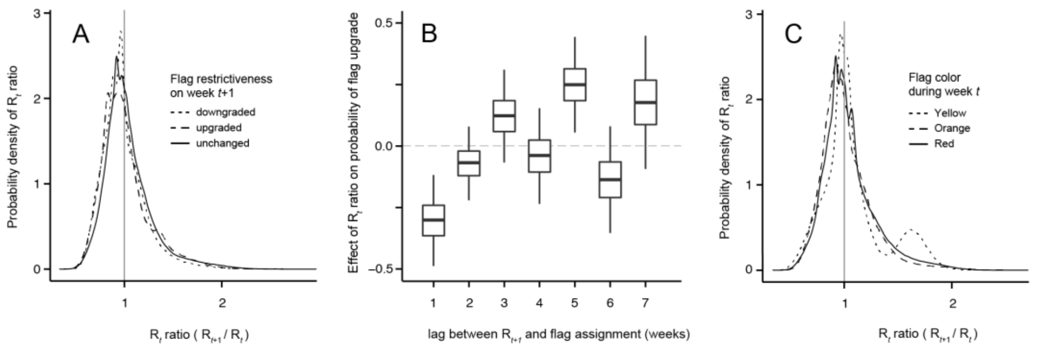

**Figure S3.** Relationship between $R_t$ and CDM flag color, or lack thereof, when running the epidemiological model with data restrict until 26 October 2020. Probability distribution of $R_t$ ratio (measuring change from week *t* to *t+1*) is the same when flags are upgraded, downgraded, or stay the same in week *t+1* (A). The probability of a flag upgrade increases with $R_t$ ratio five weeks after *t+1* (B). The probability distribution of $R_t$ ratio does not change with flag color on week *t* (C).

### Supplementary Code 1

The stan code for the epidemiological model presented here was initially developed by the Imperial College COVID-19 Response Team, with an early version published by Flaxman *et al*. (2020) *Nature* 584: 257-261. We worked with a version of the Flaxman *et al*. code kindly shared with us by Thomas Mellan and adapted it for our analysis. To run, you will need the 'Code_1.RData' file, which is available in the author's' data repository.

```
# epidemiological model
sink("base.stan")
cat("
data {
  int <lower=1> M;    // number of regions
  int <lower=1> N0;   // number of days for which to impute infections
  int <lower=1> N[M]; // days of observed data for region m. each entry must be <= N2
  int <lower=1> N2;   // days of observed data + # of days to forecast
  int cases[N2,M];   // reported cases
  int deaths[N2, M];  // reported deaths. rows with i>N contain -1. should be ignored
  matrix[N2, M] f;    // ifr
  int P;             // covariates
  matrix[N2, P] X[M];  // mobility indicators
  int EpidemicStart[M]; // 30 days before 10th death for region m
  real pop[M];         // population size
  real SI[N2];         // vector of serial interval prob dens from Flaxman et al 2020
  int W;             // number of weeks for weekly effects
  int week_index[M,N2]; // index for week effects
}

transformed data {
  vector[N2] SI_rev;   // SI in reverse order
  vector[N2] f_rev[M];  // f in reversed order
  for(i in 1:N2)
    SI_rev[i] = SI[N2-i+1];
  for(m in 1:M){
    for(i in 1:N2) {
      f_rev[m, i] = f[N2-i+1,m];
    }
  }
}

parameters {
  real<lower=0> mu[M];             // intercept for Rt
  real<lower=0> y[M];              // seed infections
  real<lower=0> phi;               // sd for expected deaths
  real<lower=0> tau;               // parameter for seed infections
  real <lower=0> ifr_noise[M];      // noise for expected deaths
  real alpha[P];                   // covariate shared between all regions m
  real alpha1[P,M];                 // covariate for each region m
  matrix[W+1,M] weekly_effect;       // weekly effect
  real<lower=0, upper=1> weekly_rho;  // weekly effect from 1 week ago
  real<lower=0, upper=1> weekly_rho1; // weekly effect from 2 weeks ago
  real<lower=0> weekly_sd;           // sd for AR
}

transformed parameters {
  matrix[N2, M] prediction = rep_matrix(0,N2,M); // for predicted cases
  matrix[N2, M] E_deaths  = rep_matrix(0,N2,M);  // for expected deaths
  matrix[N2, M] Rt = rep_matrix(0,N2,M);        // for crude Rt
  matrix[N2, M] Rt_adj = Rt;                  // for adjusted Rt
  {
    matrix[N2,M] cumm_sum = rep_matrix(0,N2,M);  // for cuml sum of predicted cases
    vector[N2] linear_effect;                 // for linear effect
    for (m in 1:M){
      linear_effect = rep_vector(0,N2);        // length of linear effect region m
      prediction[1:N0,m] = rep_vector(y[m],N0);  // learn n of cases in first N0 days
      cumm_sum[2:N0,m] = cumulative_sum(prediction[2:N0,m]);
      for(p in 1:P) {
        linear_effect -= X[m,,p] * (alpha[p] + alpha1[p,m]);
      }                       // mobility effect
      Rt[,m] = mu[m] * 2 * inv_logit((linear_effect) - weekly_effect[week_index[m],m]);            // rough Rt
```

```
      Rt_adj[1:N0,m] = Rt[1:N0,m]; // adjusted Rt until N0 is the same that rough Rt
      for (i in (N0+1):N2) { //after N0 until N2
        // scalar product of the  predicted cases by tail rev serial interval
        real convolution = dot_product(sub_col(prediction, 1, m, i-1), tail(SI_rev, i-1));
        // cumulative sum of predicted cases
        cumm_sum[i,m] = cumm_sum[i-1,m] + prediction[i-1,m];
        // infected pop decrease the rough Rt
        Rt_adj[i,m] = ((pop[m]-cumm_sum[i,m]) / pop[m]) * Rt[i,m];
        // number of predicted cases at time i for region m
        prediction[i, m] = Rt_adj[i,m] * convolution;
      }
      E_deaths[1, m]= 1e-15 * prediction[1,m];
      for (i in 2:N2){
        E_deaths[i,m] = ifr_noise[m] * dot_product(sub_col(prediction, 1, m, i-1), tail(f_rev[m], i-1));
      } // expected deaths at time i for region m
    }
  }
}

model {
  // prior distribution to parameter of seed infections
  tau ~ exponential(0.03);
  // prior distribution for standard deviation of the stationary distribution
  weekly_sd ~ normal(0,0.2); // (mu, sigma)
  // prior distribution of weekly effect from 1 week ago
  weekly_rho ~ normal(0.8, 0.05);
  //prior distribution of weekly effect from 2 week ago
  weekly_rho1 ~ normal(0.1, 0.05);
  for (m in 1:M){
    // prior distribution of seed infections in N0 days
    y[m] ~ exponential(1/tau);
    weekly_effect[3:(W+1), m] ~ normal( weekly_effect[2:W,m]* weekly_rho + weekly_effect[1:(W-1),m]* weekly_rho1,
    //weekly effect to AR process after 3rd week
    weekly_sd *sqrt(1-pow(weekly_rho,2)-pow(weekly_rho1,2) - 2 * pow(weekly_rho,2) * weekly_rho1/(1-weekly_rho1)));
  }
  weekly_effect[2, ] ~ normal(0,weekly_sd *sqrt(1-pow(weekly_rho,2)-pow(weekly_rho1,2) - 2 * pow(weekly_rho,2) *
weekly_rho1/(1-weekly_rho1)));
  // start of the weekly effect to AR process
  weekly_effect[1, ] ~ normal(0, 0.01);
  // prior distribution for death model
  phi ~ normal(0,5);
  // prior distribution for R0 in infection model
  // citation: https://academic.oup.com/jtm/article/27/2/taaa021/5735319
  mu ~ normal(3.28, 0.5);
  // prior distribution for covariate of region specific effects
  alpha ~ normal(0,0.5);
  for (i in 1:P)
  // prior distribution for covariate effects shared between regions
  alpha1[i,] ~ normal(0,0.125);
  // prior distribution for expected deaths in infection model
  ifr_noise ~ normal(1,0.1);
  for(m in 1:M){
    deaths[EpidemicStart[m]:N[m], m] ~ neg_binomial_2(E_deaths[EpidemicStart[m]:N[m], m], phi);
  } // death model
}",fill=TRUE)
sink()

# load workspace
load(“Code_1.RData”)

# load libraries
library(rstan)

# fit the model to data
# Object 'm' is of class 'stanmodel' and results from the compilation
# of the 'base.stan' code above. Object 'm' is given in the Code_1.RData
# file, along with the 'stan_data' object. For shortness, we omit the
# compilation and data formatting commands.
fit <- sampling(m, data=stan_data, iter=4000,  warmup=2000, chains=4, thin=1,
        control = list(adapt_delta = 0.99, max_treedepth = 15))
```

### Supplementary Code 2

Code for exploring relationship between CDM flags and $R_t$ estimates. Objects necessary for running this code can be found in the ‘Code_2.RData’ file, available in the author’s data repository.

```
# Load libraries
library(lubridate)
library(ggplot2)

## Load data
load("Code_2.RData")
# Controlled-Distancing Model (CDM) weeks
perdc
# Flag colors assigned to each region
# 0=yellow; 1=orange; 2=red; 3=black
bandef
# Epidemiological model results
# object fit not called for display in this code because it is too big
# this objects results from analysis carried out by Code 1
# If we were working directly with the fit object output from stan
# we would use the commands:
# out <- rstan::extract(fit)

# Here, to avoid carrying the large fit object around, we already
# have the samples from the posterior distribution of Rt in the
# out2 object, which is included in the 'Code_2.RData'
out2

## Process data
# Create array to fill with Rt values by week, region and iteration
niter <- dim(out2)[1]
nweeks <- dim(perdc)[1]
nreg <- length(covidregions)
RtWeekReg <- array(data=NA,dim=c(niter,nweeks,nreg))

# Fill the array wit Rt values
for(i in 1:length(covidregions)){
 print(i)
 N <- length(dates[[i]])
 cregion <- covidregions[[i]]
 cdates <- as_date(as.character(dates[[i]]))
 # Sets the starts of each CDM week
 starts <- which(cdates %in% perdc$start)
 # Sets the ends of each CDM week
 ends <- which(cdates %in% perdc$end)
 tends <- ends[which(ends > starts[1])]
 # Calculate the weekly Rt of region i
 for(j in 1:length(starts)) {
  # Sets the start of the week to calculate Rt
  cstarts <- starts[j]
  # Sets the end of the week to calculate Rt
  ctends <- tends[j]
  period <- tends[j]-starts[j]+1
  wstarts <- which(perdc$start==cdates[cstarts])
  # Calculate the geometric mean of the Rt for the week from daily estimates
  RtWeekReg[,wstarts,i] <- apply(out2[,cstarts:ctends,i],1,prod)^(1/period)
 } # j
} # i

# Create array to fill with ratio Rt+1/Rt by week, region and iteration
RtTransReg <- array(data=NA,dim=c(niter,nweeks,nreg))

# Fill the array wit Rt ratios
for(i in 1:length(covidregions)){
 print(i)
 # Search first week which will give numerator Rt for ratio
 numwk <- which(!is.na(RtWeekReg[1,,i]))[2]
 for (j in numwk:nweeks){
  # Calculate Rt+1/Rt ratio and store in RtTransReg
  RtTransReg[,numwk,i] <- RtWeekReg[,numwk,i]/RtWeekReg[,numwk-1,i]
```

```
    RtTransReg[,j,i] <- RtWeekReg[,j,i] / RtWeekReg[,(j-1),i]
    #RtTransReg[,(numwk-1),i] <- RtWeekReg[,numwk,i]/RtWeekReg[,numwk-1,i]
  } # j
} # i

# Remove the first column without data
RtTransReg <- RtTransReg[,-1,]

##Figure 2A

#Identify transitions with flags changes
tbandef <- as.matrix(bandef[,2:ncol(bandef)])
dflags1 <- t(tbandef[,2:ncol(tbandef)]-tbandef[,1:(nweeks-1)])

# Create vectors to fill with probability density of Rt ratio
# according to flag changes
ndown1 <- dim(which(dflags1<0,arr.ind = TRUE))[1]
nunchan1 <- dim(which(dflags1==0,arr.ind = TRUE))[1]
nup1 <- dim(which(dflags1>0,arr.ind = TRUE))[1]
RtTransdown1 <- rep(NA,niter*ndown1)
RtTransunchan1 <- rep(NA,niter*nunchan1)
RtTransup1 <- rep(NA,niter*nup1)

# Fill vectors
for(i in 1:niter) {
  citer1 <- RtTransReg[i,,]
  RtTransdown1[(i*ndown1-ndown1+1):(i*ndown1)] <- citer1[which(dflags1<0)]
  RtTransunchan1[(i*nunchan1-nunchan1+1):(i*nunchan1)] <- citer1[which(dflags1==0)]
  RtTransup1[(i*nup1-nup1+1):(i*nup1)] <- citer1[which(dflags1>0)]
}

# Create vector with values only for regions and weeks with estimates
Rtdown1 <- RtTransdown1[which(!is.na(RtTransdown1))]
Rtunchan1 <- RtTransunchan1[which(!is.na(RtTransunchan1))]
Rtup1 <- RtTransup1[which(!is.na(RtTransup1))]

# Put vectors in dataframe
dfrts <- data.frame(
  rts1 = c(Rtdown1, Rtunchan1,Rtup1),
  flags1 = c(rep("downgraded",length(Rtdown1)),rep("unchanged",length(Rtunchan1)),rep("upgraded",length(Rtup1)))
)

# Ploting
ggplot(dfrts) +
  geom_density(aes(rts1,linetype=flags1)) +
  ylab("Probability density of Rt ratio") +
  xlab("Rt ratio (Rt+1/Rt)") +
  theme_classic() +
  guides(linetype = guide_legend(title = "Flag restrictiveness on week t+1")) +
  geom_vline(xintercept = 1)

## Figure 2C

# Create a matrix with flags
cdmf <- t(as.matrix(bandef[,2:ncol(bandef)-1]))

# Identify frequency of each flag
nyellow <- dim(which(cdmf==0,arr.ind = TRUE))[1]
norange <- dim(which(cdmf==1,arr.ind = TRUE))[1]
nred <- dim(which(cdmf==2,arr.ind = TRUE))[1]
nblack <- dim(which(cdmf==3,arr.ind = TRUE))[1]

# Create vectors to fill with probability density of Rt ratio according to flag colors
RtTransnyellow <- rep(NA,niter*nyellow)
RtTransnorange <- rep(NA,niter*norange)
RtTransnred <- rep(NA,niter*nred)
RtTransnblack <- rep(NA,niter*nblack)

# Fill vectors
for(i in 1:niter) {
  citer1 <- RtTransReg[i,,]
```

```
  RtTransnyellow[(i*nyellow-nyellow+1):(i*nyellow)] <- citer1[which(cdmf==0)]
  RtTransnorange[(i*norange-norange+1):(i*norange)] <- citer1[which(cdmf==1)]
  RtTransnred[(i*nred-nred+1):(i*nred)] <- citer1[which(cdmf==2)]
  #RtTransnblack[(i*nblack-nblack+1):(i*nblack)] <- citer1[which(cdmf==3)]
}

# Create vector with values only for regions and weeks with estimates
Rtfyellow <- RtTransnyellow[which(!is.na(RtTransnyellow))]
Rtforange <- RtTransnorange[which(!is.na(RtTransnorange))]
Rtfred <- RtTransnred[which(!is.na(RtTransnred))]
Rtfblack <- RtTransnblack[which(!is.na(RtTransnblack))]

# Put vectors in dataframe
dfbsrt <- data.frame(
  rts1 = c(Rtfyellow, Rtforange,Rtfred),
  flags1 = c(rep("Yellow",length(Rtfyellow)),rep("Orange",length(Rtforange)),rep("Red",length(Rtfred)))
)

# Plotting
ggplot(dfbsrt) +
  geom_density(aes(rts1,linetype=flags1)) +
  ylab("Probability density of Rt ratio") +
  xlab("Rt ratio (Rt+1/Rt)") +
  theme_classic() +
  guides(linetype = guide_legend(title = "Flag color during week t")) +
  geom_vline(xintercept = 1)

## Figure 2B
# Standardize Rt ratios for linear model
RtTrRgSt<- (RtTransReg-mean(RtTransReg,na.rm=TRUE)) / sd(RtTransReg,na.rm=TRUE)
# Create a time-by-region matrix of flag upgrade (1) or no upgrade (0)
bandup <- t((bandef[,3:33]-bandef[,2:32]) > 0)

# Define the lag between Rt ratio and flag upgrade,
# change the value for longer lags
lag <- 1
# Create a Rt ratio array that matches the intended lag
Rtratios <- RtTrRgSt[,1:(dim(RtTrRgSt)[2]-lag),]
# Create a flag upgrade matrix that also matches the lag
BandUp <- bandup[(1+lag):31,]

# Fit a logistic regression to the effect of Rt ratio on probability of
# flag upgrade after the given lag
logregcoef <- rep(NA,niter) # prepare vector for storing coefficients
for(i in 1:niter) { # fits one regression per iteration
  print(i)
  # Extract vectors from Rt ratios correspondents to flag upgrade or not flag upgrade
  Rtrbandup <- Rtratios[i,,][which(BandUp)] # select Rt ratios
  Rtrbandnup <- Rtratios[i,,][which(!BandUp)]
  Rtrbandup <- Rtrbandup[which(!is.na(Rtrbandup))] # remove nas
  Rtrbandnup <- Rtrbandnup[which(!is.na(Rtrbandnup))]
  # Create a dataframe for these vectors
  Rtt <- c(Rtrbandup,Rtrbandnup)
  bdt <- c(rep(1,length(Rtrbandup)),rep(0,length(Rtrbandnup)))
  glm.dados <- data.frame(Rtt,bdt)
  # Estimate the effect by GLM
  out.glm <- glm(bdt~Rtt,family=binomial,data=glm.dados)
  # Save the coefficient for each iteration i
  logregcoef[i] <- out.glm$coefficients[2]
}

# Save the vector with coefficients for this lag
d1logregcoef <- logregcoef

# Create a dataframe with coefficients from all lags
rtsobreab <- data.frame(
  efeito =
c(d1logregcoef,d2logregcoef,d3logregcoef,d4logregcoef,d5logregcoef,d6logregcoef,d7logregcoef,d8logregcoef,d9logregcoef,d
10logregcoef),
  delay = c(rep(1, 8000),rep(2, 8000),rep(3, 8000),rep(4, 8000),rep(5, 8000),rep(6, 8000),rep(7, 8000),rep(8, 8000),rep(9,
8000),rep(10, 8000))
```

```
)

# Plotting
ggplot(rtsobreab) +
  geom_boxplot(aes(x=delay, y=efeito,  group=delay), coef=1, outlier.shape = NA) +
  labs(x="Lag", y = "Rt+1/Rt effect") +
  scale_y_continuous(limits = c(-0.5,0.5)) +
  scale_x_continuous(breaks = seq(from = 1, to = 10, by = 1)) +
  theme_classic() +
  geom_hline(yintercept=0, linetype="dashed", size=0.5)
```

### Supplementary Code 3

The following code implements a mixed-effects model of the variation in human mobility according to CDM flag color, time, and COVID region. We fit the same model to six different Google mobility metrics, which are called, one at a time, at the beginning of the code. The analysis was implemented in R with model description and MCMC sampling in jags. The code is adapted from Marc Kéry's (2010) *Introduction to WinBUGS for Ecologists*, Chapter 12, pages 151-166. To run this in R you can obtain the 'Code_3.RData' file from the data repository. The RData file contains all the necessary data.

```
# load workspace
load("Code_3.RData")

# load libraries
library(jagsUI)

# choose the google metric to load
vimob <- mobilidaderegiao_bandeiras$residencias # residences
#vimob <- mobilidaderegiao_bandeiras$comercios  # retail and recreation
#vimob <- mobilidaderegiao_bandeiras$mercados  # grocery and pharmacies
#vimob <- mobilidaderegiao_bandeiras$parques   # parks
#vimob <- mobilidaderegiao_bandeiras$estacoes  # transit stations
#vimob <- mobilidaderegiao_bandeiras$trabalho  # workplace

# Mixed-effects mobility model
# Model definition
sink("lme.model2.txt")
cat("
model {

 # Priors
 for (i in 1:4){
  alpha[i] ~ dnorm(0, 0.001)                  # Fixed intercepts
 }
 for (i in 1:ngroups){
  beta[i] ~ dnorm(mu.slope, tau.slope)        # Random slopes
 }
 for (i in 1:225){
  beta1[i] ~ dnorm(mu.slope1, tau.slope1)     # Random slopes1
 }

 mu.slope ~ dnorm(0, 0.001)                   # Mean hyperparameter for random slopes
 tau.slope <- 1 / (sigma.slope * sigma.slope)
 sigma.slope ~ dunif(0, 100)                  # SD hyperparameter for slopes
 mu.slope1 ~ dnorm(0, 0.001)                  # Mean hyperparameter for random slopes1
 tau.slope1 <- 1 / (sigma.slope1 * sigma.slope1)
 sigma.slope1 ~ dunif(0, 100)                 # SD hyperparameter for slopes1
 tau <- 1 / ( sigma * sigma)                  # Residual precision
 sigma ~ dunif(0, 100)                        # Residual standard deviation

 # Likelihood
 for (i in 1:n) {
  vimob[i] ~ dnorm(mu[i], tau)
  mu[i] <- alpha[iband[i]] + beta[regiao[i]] + beta1[itemp[i]]
 }
}
",fill=TRUE)
sink()

# run the mixed-effects mobility model
out <- jags(win.data, inits, parameters, "lme.model2.txt", n.thin=nt, n.chains=nc, n.burnin=nb, n.iter=ni)
```

### Supplementary Code 4

Code for obtaining infection fatality ratios for RS COVID regions based on demographic data. Computation was implemented in R using code developed by the Imperial College COVID-19 Response Team for their COVID-19 Report 21 and kindly shared with us by Thomas Mellan. To run, you will need the ‘Code_4.RData’ file, available in the data repository.

```
# load workspace
load(“Code_4.RData”)

# load libraries
library(dplyr)
library(magrittr)
library(tidyverse)
library(readxl)
library(socialmixr)
library(dfoptim)

# compute ifr for each COVID region
for (i in 1:length(covidreg)){

 raw_covidregion_pop <- unlist(demog[demog$regiaocovid == covidreg[i], pop_columns])
 covidregion_pop <- c(raw_covidregion_pop[1:13], sum(raw_covidregion_pop[14:17]))

 # Processing the Contact Matrix to Generate a Probability Matrix
 # (how much a person in each age group will mix with people
 # from a different age group)
 MIJ <- t(sapply(seq(covidregion_pop),function(x){
  contact_mat[x,]*covidregion_pop[x]
 }))
 adjust_mat<-(MIJ+t(MIJ))/2
 new_mix_mat<-t(sapply(seq(covidregion_pop),function(x){
  adjust_mat[x,]/covidregion_pop[x]
 }))
 c_mat<-t(sapply(seq(covidregion_pop),function(x){
  new_mix_mat[x,]/sum(new_mix_mat[x,])
 }))

 ai <- rowSums(new_mix_mat)
 ng_eigen <- Re(eigen(new_mix_mat)$val[1])
 rmod <- R/ng_eigen*ai
 tot_pop <- sum(covidregion_pop)

 # Running an Optimiser to Get the Number Infected by Age for Each Age Group
 x <- get_AR(R = R, rmod = rmod, c_mat = c_mat, demog = covidregion_pop, init = init, guess_hom = guess_hom,
                    guess_modifiers = guess_modifier, iterates = iterates, reiterates = reiterates, restarts = restarts, tol = tol)

 # Number infected and attack rate for 5 year age bands up to 75+
 number_inf_by_age <- x$par
 attack_rates_by_age <- x$par/covidregion_pop

 # Splitting up 75+ into 75-80 and 80+ to incorporate the age-specific IFRs
 # for these two groups that we have
 infs_65_70 <- number_inf_by_age[14] * raw_covidregion_pop[14]/(sum(raw_covidregion_pop[14:17]))
 infs_70_75 <- number_inf_by_age[14] * raw_covidregion_pop[15]/(sum(raw_covidregion_pop[14:17]))
 infs_75_80 <- number_inf_by_age[14] * raw_covidregion_pop[16]/(sum(raw_covidregion_pop[14:17]))
 infs_80_plus <- number_inf_by_age[14] * sum(raw_covidregion_pop[17])/(sum(raw_covidregion_pop[14:17]))
 number_inf_by_age[14] <- infs_65_70
 number_inf_by_age[15] <- infs_70_75
 number_inf_by_age[16] <- infs_75_80
 number_inf_by_age[17] <- infs_80_plus

 # Calculating the number of Deaths in Each Age Group
 deaths <- number_inf_by_age * IFRs
 IFR[i] <- sum(deaths)/sum(number_inf_by_age)
}

#return IFRs
data.frame("covidregion" = covidreg, "IFR" = IFR)
```
